# Genomic surveillance, characterisation and intervention of a carbapenem-resistant *Acinetobacter baumannii* outbreak in critical care

**DOI:** 10.1101/2020.08.10.20166652

**Authors:** Leah W. Roberts, Brian M. Forde, Trish Hurst, Weiping Ling, Graeme R. Nimmo, Haakon Bergh, Narelle George, Krispin Hajkowicz, John F McNamara, Jeffrey Lipman, Budi Permana, Mark A. Schembri, David Paterson, Scott A. Beatson, Patrick N. A. Harris

## Abstract

Infections caused by carbapenem-resistant *Acinetobacter baumannii* (CR-Ab) have become increasingly prevalent in clinical settings and often result in significant morbidity and mortality due to their multidrug resistance (MDR). Here we present an integrated whole genome sequencing (WGS) response to a polymicrobial outbreak in a Brisbane hospital between 2016-2018. 28 CR-Ab (and 21 other MDR Gram negative bacilli) were collected from Intensive Care Unit and Burns Unit patients and sent for WGS with a 7-day turn-around-time in clinical reporting. All CR-Ab were sequence type (ST)1050 and within 10 single nucleotide polymorphisms (SNPs) apart, indicative of an ongoing outbreak, and distinct from historical CR-Ab isolates from the same hospital. Possible transmission routes between patients were identified on the basis of CR-Ab and *K. pneumoniae* SNP profiles. Continued WGS surveillance between 2016 to 2018 enabled suspected outbreak cases to be refuted, but a resurgence of the outbreak CR-Ab mid-2018 in the Burns Unit prompted additional screening. Environmental metagenomic sequencing identified the hospital plumbing as a potential source. Replacement of the plumbing and routine drain maintenance resulted in rapid resolution of the secondary outbreak and significant risk reduction with no discernable transmission in the Burns Unit since. Here we demonstrate implementation of a comprehensive WGS and metagenomics investigation that resolved a persistent CR-Ab outbreak in a critical care setting.

## Introduction

Hospital outbreaks of multi-drug resistant Gram-negative pathogens present great risk to patients and are costly(*1, 2*). Whole genome sequencing (WGS) has been proposed as an effective tool to support infection control responses to emerging outbreaks within the healthcare environment, but barriers exist to the effective implementation into clinical practice(*3*).

*Acinetobacter baumannii* has emerged over recent decades as a major nosocomial pathogen(*4*). Its capacity to develop or acquire resistance to multiple antibiotic classes, in addition to intrinsic resistance to desiccation and disinfectants, contributes to persistence of *A. baumannii* in the hospital environment(*5*, *6*). It has frequently been a cause of nosocomial outbreaks, particularly in the critical care setting(*7*-*9*). *A. baumannii* are often resistant to multiple antibiotic classes and the global incidence of extensively-drug resistant (XDR) or even pan-drug resistant (PDR) strains has been increasing(*10*-*12*). Carbapenem-resistant *A. baumannii* (CR-Ab) have been seen at high prevalence in several areas, particularly in the Asian-Pacific region, Latin America and the Mediterranean(*13*). Carbapenem resistance in *A. baumannii* usually arises from the acquisition of genes encoding carbapenemases, particularly OXA-type carbapenemases (e.g. OXA-23), and may be associated with high mortality in vulnerable patients(*14*).

Here we describe a large outbreak of CR-Ab, and other co-infecting MDR Gram-negative pathogens, occurring within an Intensive Care Unit (ICU) and burns facility. Incorporation of whole genome sequencing (WGS) in real-time facilitated rapid characterisation of this complex polymicrobial outbreak, provided a detailed understanding of transmission pathways and helped to direct a successful infection control response.

## Case study

A 25-year old patient with extensive burn injuries was retrieved from an overseas healthcare facility. As per infection control protocols, the patient was placed on contact precautions and provided a single room. Initial nasal and rectal screening swabs were negative for MDR pathogens, including CR-Ab. An extended-spectrum beta-lactamase (ESBL)-producing *Klebsiella pneumoniae* was isolated from the patient’s respiratory secretions on day 4, and within 24-hours a similar organism was isolated from blood cultures. Repeated collection of blood cultures demonstrated a polymicrobial culture with ESBL-producing *K. pneumoniae*, CR-Ab and *Pseudomonas aeruginosa* on day 6, that tested susceptible to all first line agents. Over the following days, CR-Ab was also isolated from numerous clinical specimens, including a femoral line tip, endotracheal aspirates, rectal swabs, wound swabs and operative specimens collected from debrided tissue. Blood cultures repeatedly grew CR-Ab, (day 15 and 45 of admission), with the emergence of colistin resistance when tested by Etest (MIC 32 µg.mL^-1^) on day 45. *Serratia marcescens* was co-cultured in blood on day 15 and was also grown from respiratory secretions and wounds swabs.

Over the next 5 months in 2016, 18 additional patients within the same Intensive Care Unit (ICU) area were also found to be colonized or infected with phenotypically similar CR-Ab, *K. pneumoniae, S. marcescens* and/or *P. aeruginosa*. This included CR-Ab colonized cases identified in patients discharged from the ICU to the Burns Unit or other surgical wards throughout the hospital, and eventually patients admitted to the Burns Unit. The final CR-Ab case was identified several weeks later in a patient discharged from the Burns Unit and transferred to a hospital in a remote part of Queensland. An outbreak investigation team was constituted as soon as it was suspected that an outbreak of CR-Ab had occurred within the ICU and the use of WGS for strain characterization was initiated.

## Results

### WGS predicted likely transmission pathways and ruled out non-outbreak cases

Between May to August 2016, a total of 55 isolates were recovered from 22 patients (see supplementary data 1). These isolates included *A. baumannii, K. pneumoniae, S. marcescens, E. cloacae* and *P. aeruginosa*. Species typing and antibiogram analysis alone were insufficient to determine clonal relationships between these isolates. As such, we used WGS to establish the relationship between isolates and predict patient transmission based on SNP accumulation.

We applied WGS in real-time over the course of the outbreak. Four reports aimed at communicating genomic analyses to infection control and other clinical staff at RBWH were delivered during the primary outbreak (June 22, July 15, Aug 2 and Aug 29). We managed on average a one-week turn-around time between receiving the isolates and presenting a finalised report, which consistent of [i] a front page overview of the analysis and key outcomes/interpretations conveyed as short bullet points, [ii] detailed analysis and diagrams on the internal pages, and [iii] method descriptions (see supplementary methods and results 1). Actual time between receipt of sequencing data and reporting was 8-72 hours depending on the complexity of analyses with supplementary interim reports and regular academic-clinical partner meetings necessary to communicate our comparative genomic analyses and help shape the content of the final reports (see supplementary reports 1 and 2 for example reports from June 22 and Aug 29, respectively).

The presumed index patient admitted in early May 2016 was identified with ST1050 CR-Ab, ST515 *K. pneumoniae*, ST979 *P. aeruginosa* and *S. marcescens*. Using WGS, we found that 16 of the 21 patients admitted following the index patient had bacterial infections related to either the ST1050 CR-Ab or the ST515 *K. pneumoniae*. Transmission direction based on the accumulation of SNPs was inferred in patients 10, 11, 13, 14, 15, 16 and 17 (Figure 1A, as indicated by lines with arrows). CR-Ab isolates from the first 9 patients (and patient 12) were identical based on core SNPs, making inference of patient transmission impossible using SNPs alone. However, when combined with SNP information from *K. pneumoniae* isolates, it was possible to infer co-transmission of *K. pneumoniae* and CR-Ab from the index patient to patient 6 (Figure 1 and supplementary methods and results 1).

**Figure 1:**
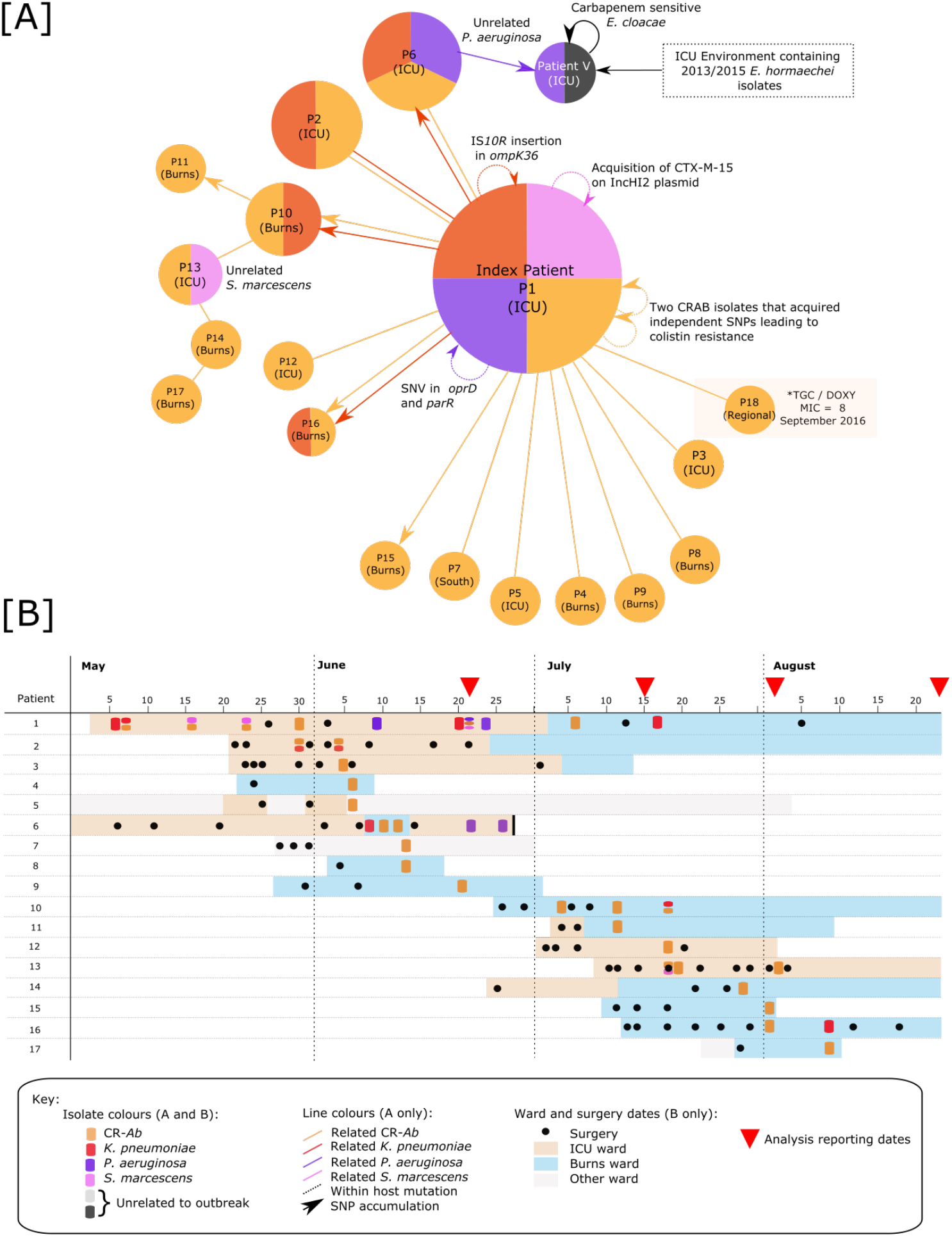
Patient relationship matrix describing 2016 outbreak of CR-Ab. [A] Each circle represents a patient, where the size of the circle correlates to the number of isolates from that patient. Colours correspond to bacterial species. Straight lines connecting circles represent patients with identical isolates at the core genome level (and as such directionality of transmission cannot be inferred). Lines with arrows (coloured by species) represent predicted direction of transmission based on the accumulation of SNPs between patients’ isolates. Circular arrows represent changes in individual patient’s isolates, [B] timeline of patient samples, as well as location and surgery dates.

Strains of *S. marcescens* and *P. aeruginosa* specific to the index patient were not found in other patients (see supplementary material). Two patients had unrelated *S. marcescens* and *P. aeruginosa* isolates (patients 13 and 6, respectively). Transmission of the unrelated *P. aeruginosa* isolate from patient 6 to another patient in the ICU ward (denoted patient V) was detected. Patient V was also found to have an *Enterobacter cloacae* isolate (later identified as *Enterobacter hormaechei* by WGS) identical to that identified in a 2015 outbreak from the same hospital(*15*). This patient also carried an additional carbapenem-sensitive *E. cloacae* (bla_IMP-4_ negative) that was unrelated to the carbapenem-resistant isolate.

Over the course of the outbreak, each species carried by the index patient acquired additional antibiotic resistance mechanisms, via mutations or plasmid gain (Figure 1 and supplementary methods and results 1).

An additional CR-Ab was isolated in September 2016 from a patient in a Regional Queensland (QLD) hospital who had previously been admitted to the Brisbane ICU (patient 18, isolate MS14438). Analysis of this isolate found that it was closely related to isolates from the initial outbreak between May to August 2016.

Extensive environmental swabbing throughout the ICU and Burns Unit was conducted on the 16^th^ of June 2016, targeting patient bedrooms as well as high-touch areas (e.g. Nurse keyboards, trolley, door handles). However, no bacterial species related to the CR-Ab outbreak were detected in the environment based on traditional culture methods using chromogenic agar.

### The outbreak CR-Ab was likely imported into the hospital ICU

29 CR-Ab isolates related to the ongoing outbreak were collected from 18 patients Between May-September 2016. All were found to be ST1050 (global clone [GC] 2) and less than 10 SNPs different (fig. S2). Three carbapenem-sensitive *A. baumannii* isolated at the same time were found to be different sequence types and unrelated to the outbreak. Comparison of the outbreak ST1050 CR-Ab isolates to historical CR-Ab isolates collected between 2000-2016 from the hospital found no close relationship, indicating that the CR-Ab had likely been introduced into the hospital with the index patient (fig. S3).

All ST1050 CR-Ab isolates related to the index were found to be extensively resistant to carbapenems, β-lactams, cephalosporins, aminoglycosides, and quinolones (Table 1). Resistance to colistin appeared in three isolates from the index patient and was mediated by two independent SNP acquisitions in the sensor kinase gene *pmrB* (causing the amino acid changes T235I in MS14413 and its descendant MS14402, and R263C in MS14407). Antibiotic resistance genes were conserved between all isolates, and included β-lactamases (such as *bla*_OXA-23_ and *bla*_OXA-66_), streptomycin resistance genes (*strA* and *strB*), and aminoglycoside resistance genes *(aph(3’)-Ic, aadA1* and the methylase *armA*). Finally, a single SNP was found to result in the reversion of a nonsense mutation in a putative type 3 filamentous fimbriae gene (*filB*). This SNP was identified in the majority of CR-Ab isolates taken after the 4^th^ of July 2016 and appears to have arisen independently multiple times in the *A. baumannii* lineage (supplementary methods and fig. S4).

**Table 1:** CR-Ab MICs and AB resistance genes. Table only shows select representative isolates as all CR-Ab were found to have the same AB resistance gene profile and MIC data. Colours represent mechanism of detection: blue = Etest MIC, Green = Disk diffusion zone diameter, Orange = Vitek2, Grey = Resfinder (accessed Aug 2017).

| Strain |  | MS8413 | MS8419 | MS8436 | MS8442 | MS8441 |
| --- | --- | --- | --- | --- | --- | --- |
| Patient |  | 4 | 5 | 6 | 7 | 8 |
| Site |  | Leg wound | ETA | Tissue buttock | Wound Swab | Rectal Swab |
| Colistin | Colistin | 0.125 | 0.25 | 0.25 | 0.125 | 0.5 |
| Carbapenem | Mero | 32 | >32 | >32 | >32 | >32 |
|  | Imi | >32 | >32 | n.t | >32 | n.t |
|  | Erta | >32 | >32 | >32 | >32 | >32 |
| Beta-lactam and Cephalosporins | Sulb | 32 | 32 | 64 | 32 | 64 |
|  | MER | R | R | R | R | R |
|  | TIM | R | R | R | R | R |
|  | TAZ | R | R | R | R | R |
|  | CRO | R | R | R | R | R |
|  | CAZ | R | R | R | R | R |
|  | FEP | R | R | R | R | R |
|  | KZ | R | R | R | R | R |
|  | Azt | 6mm R | 6mm R | n.t | 6mm R | n.t |
|  | CTZ/TAZ | >256 | >256 | 128 | 16 | 96 |
|  | CAZ/AVI | 16mm R | 17mm R | 18mm R | 15mm R | 18mm R |
|  | blaADC-25 | + | + | + | + | + |
|  | blaOXA-23 | + | + | + | + | + |
|  | blaOXA-66 | + | + | + | + | + |
| Aminoglycosides | Amikacin | >256 | >256 | >256 | >256 | >256 |
|  | GENT | R | R | R | R | R |
|  | TOB | R | R | R | R | R |
|  | Aph(3')-Ic-1 | + | + | + | + | + |
|  | aadA1 | + | + | + | + | + |
|  | armA | + | + | + | + | + |
| Quinolones | CIP | R | R | R | R | R |
|  | NOR | R | R | R | R | R |
| Trimethoprim/<br>Sulphonamide | TMP | R | R | R | R | R |
|  | SXT | R | R | R | R | R |
|  | Sul1 | + | + | + | + | + |
|  | Sul2 | + | + | + | + | + |
| Tigecycline | Tige | 2 | 2 | 2 | 2 | 4 |
| Chloramphenicol | Chloro | 6mm R | 6mm R | n.t | 6mm R | n.t |
|  | catB8 | + | + | + | + | + |
| Fosfomycin | Fosfo | 256 | 512 | n.t | 128 | n.t |
| Tetracycline | Doxy | 4 | 4 | 2 | 2 | 2 |
| Macrolides | mph(E)_3 | + | + | + | + | + |
|  | msr(E)_4 | + | + | + | + | + |
| Streptomycin | strA | + | + | + | + | + |
|  | strB | + | + | + | + | + |

n.t = not tested
Mero = Meropenem, Tige = Tigecycline, Sulb = Sulbactam, CTZ/TAZ = Ceftolozane/tazobactam,
CAZ/AVI = Ceftazidime/avibactam, Chloro = Chloramphenicol, Fosfo = Fosfomycin, Azt =
Aztreonam, Erta = Ertapenem, Doxy = Doxycycline, Imi = Imipenem, KZ = Cephazolin, TMP =
Trimethoprim, SXT = Co-trimoxazole, GENT = Gentamicin, TOB = Tobramycin, CRO = Ceftriaxone,
CAZ = Ceftazidime, FEP = Cefepime, TAZ = Piperacillin/tazobactam, CIP = Ciprofloxacin, NOR =
Norfloxacin, MER = Meropenem, TIM = Ticarcillin/clavulanate

### PacBio sequencing of CR-Ab reveals context of resistance genes and mobile elements

Complete sequencing of a reference CR-Ab isolate (MS14413) from the index patient using long-read sequencing provided a high-quality reference and allowed contextualization of the antibiotic resistance genes (as well as other mobile genetic elements) within the genome. Assembly of the ST1050 CR-Ab reference genome revealed a 4,082,498 bp chromosome with no plasmids. *strA, strB* and *sul2* resided within a novel AbGRI1 resistance island most closely related to the *A. baumannii* strain CBA7 (GenBank:NZ_CP020586.1) isolated from Korea in 2017 (fig. S5). The CR-Ab isolates also carried Tn6279 (also known as AbGRI3-2), which encompassed a large number of resistance genes including *mph(E)* and *msr(E)* (macrolide resistance) and the methylase gene *armA* (gentamicin resistance) (fig. S6). Resistance to carbapenems in these CR-Ab isolates was likely driven by the presence of two copies of *bla*_OXA-23_ residing in separate Tn2006 transposons within the chromosome. An *ISAba1* insertion sequence upstream of the chromosomal *ampC* gene was also detected, which has previously been shown to enhance cephalosporin resistance(*16*).

Long read sequencing also revealed a KL12 capsule (K) locus, which shares 97% nucleotide identity to the capsule region found in the GC1 *A. baumannii* strain D36 (GenBank:NZ_CP012952.1) (fig. S7). However, the *wzy* gene (a polymerase required for capsular polysaccharide biosynthesis) within the capsule locus was interrupted by an IS*Aba125* insertion sequence in all CR-Ab isolates. Further comparative analysis found a portion of the capsule locus in MS14413 to share 99% nucleotide identity to the capsule from *A. baumannii* strain BAL_097 (GenBank: KX712116), which carries a *wzy* gene at the beginning of the capsule region. This unusual gene placement also appears in MS14413, and likely complements the loss of the internal *wzy* gene (Figure 2). The high nucleotide identity at this region also indicates possible recombination. Overlapping the capsule (K) region in MS14413 (and some other patient 1 CR-Ab isolates) is a large 41,375 kb tandem duplication, encompassing *Tn2006* (resulting in 3 chromosomal copies of this transposon) and is further discussed in the Supplementary results (Figure 2).

**Figure 2:**
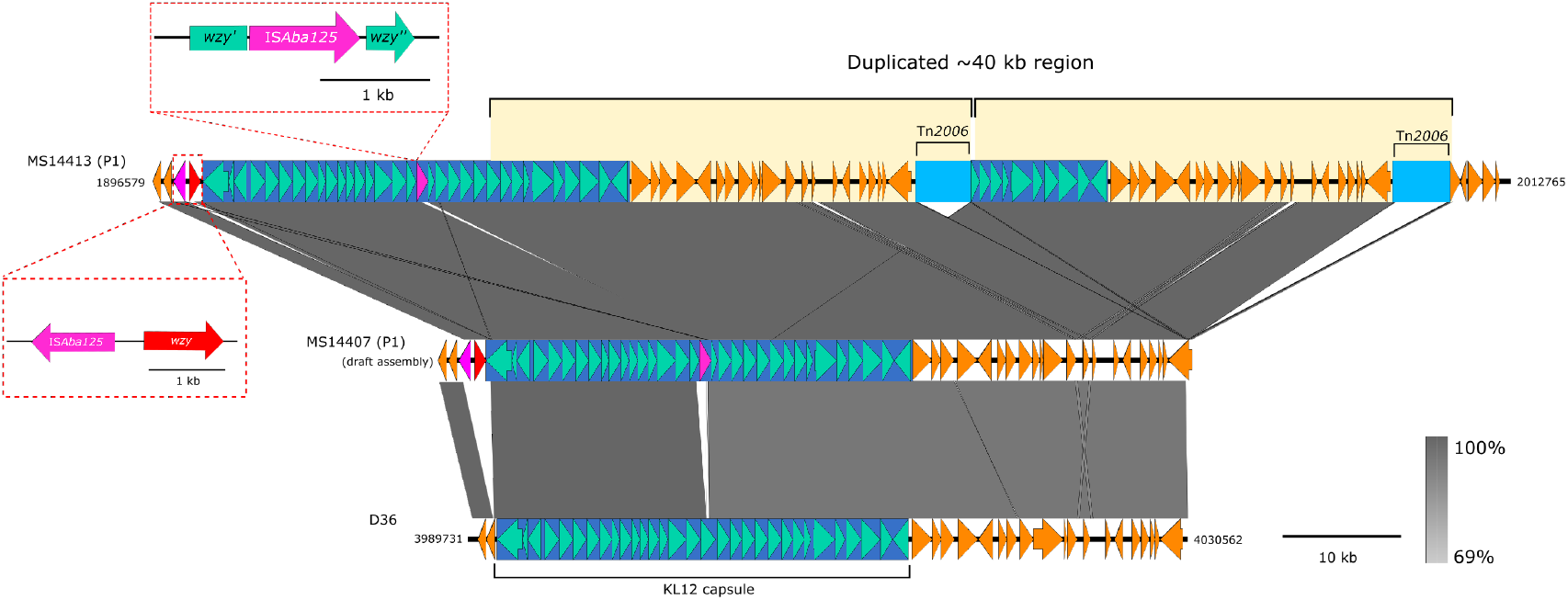
large ~41 kb tandem duplication found in MS14413. Duplication of part of the capsule (K) region in the MS14413 complete genome (top line), resulting in 3 chromosomal copies of Tn*2006* (third copy at alternate locus). This duplication appears to have arisen in some of the index patient isolates, but not other isolates involved in the outbreak (e.g. MS14407 concatenated draft genome, central line). The *wzy* gene in the capsule region was found to be interrupted by an IS*Aba125* element, however a secondary *wzy* gene was identified at the start of the capsule region. Neither the IS*Aba125* insertion or secondary *wzy* gene is found in the KL12 capsule locus of *A. baumannii* strain D36 (bottom line).

### Whole genome shotgun metagenomics detects CR-Ab in hospital environment

Ongoing surveillance was conducted using WGS following the initial outbreak. Despite continual environmental cleaning and routine swabbing, the outbreak CR-Ab strain persisted through to September 2018 (Figure 3). Swabs collected from surfaces within the ICU and Burns Unit (e.g. handles, tables, shelves, computer equipment) in 2016 and 2017 were unable to detect CR-Ab in the environment and did not yield enough DNA for direct metagenomic sequencing (data not shown).

**Figure 3:**
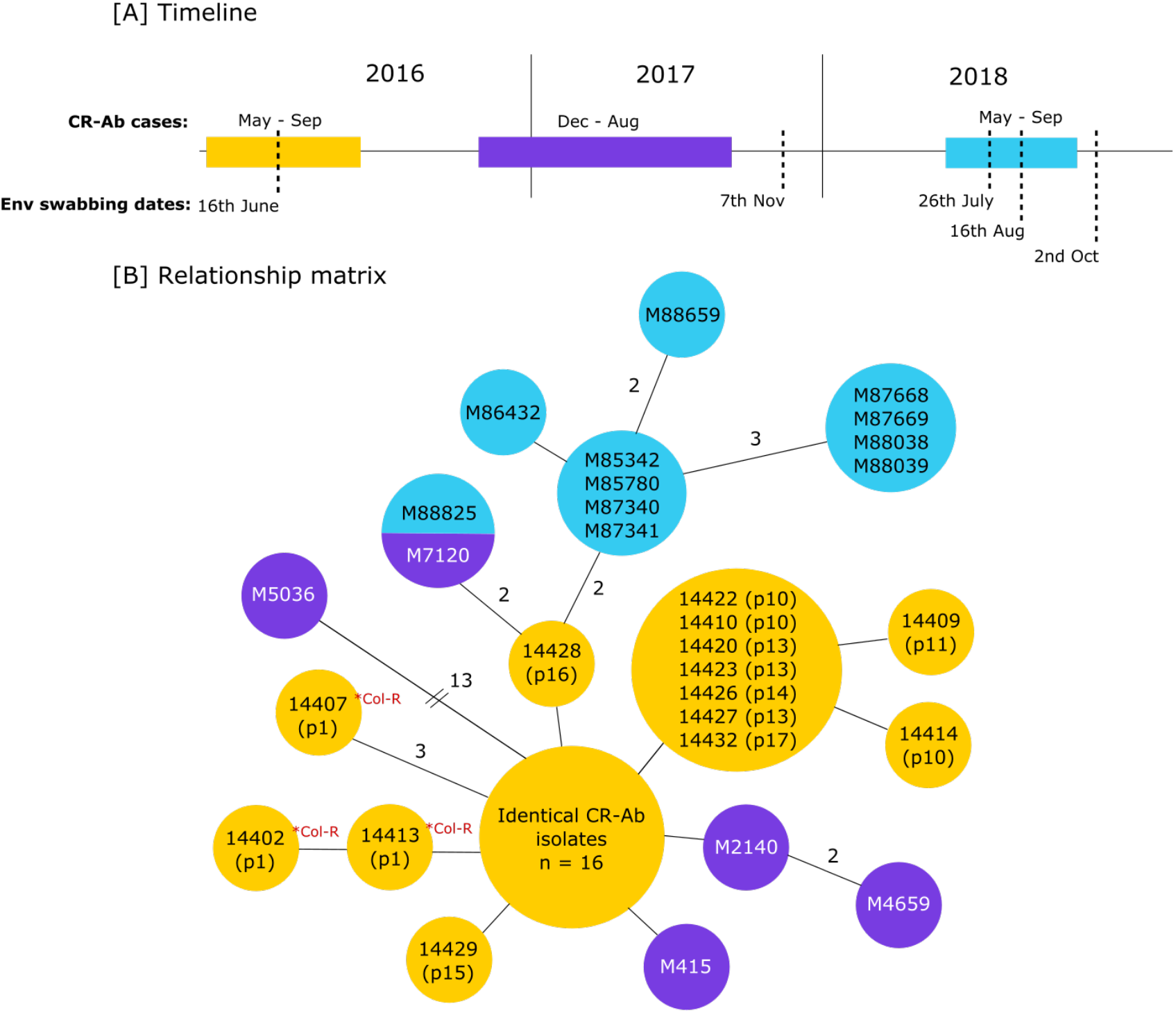
Ongoing CR-Ab surveillance from 2016-2018. [A] timeline of CR-Ab cases and dates of environmental swabbing between 2016-2018 [B] Relationship matrix of all CR-Ab isolates related to the initial outbreak. Col-R = predicted colistin resistance via mutation in *pmrB*. Isolates within the same circle are identical at the core genome. Branches represent 1 SNP difference (except where specified). Isolates from the original 2016 outbreak are in yellow. Purple isolates were collected in late 2016-2017. Isolates in blue were collected in 2018. Isolate M88825 was isolated from an Antechamber environment in 2018 and found to be identical at the core SNP level to M7120, isolated in August 2017.

Due to 11 new cases of CR-Ab detected between May to September 2018, additional environmental sampling was carried out in the Burns ward environment. Between July to October 2018, areas of presumed high bacterial load (such as floor drains, plumbing, inside burns bath drains, etc.) were targeting for environmental sampling (figure 4). All samples were subjected to culture using traditional methods (on chromogenic media) and direct DNA extraction and shotgun metagenomic sequencing. Of 50 environmental samples, two were culture positive for CR-Ab (R5666 and R5864), while four were positive based on analysis of the metagenomic sequencing data (R5515, R5510, R5863 and R5864) (table S4, fig. S11).

**Figure 4:**
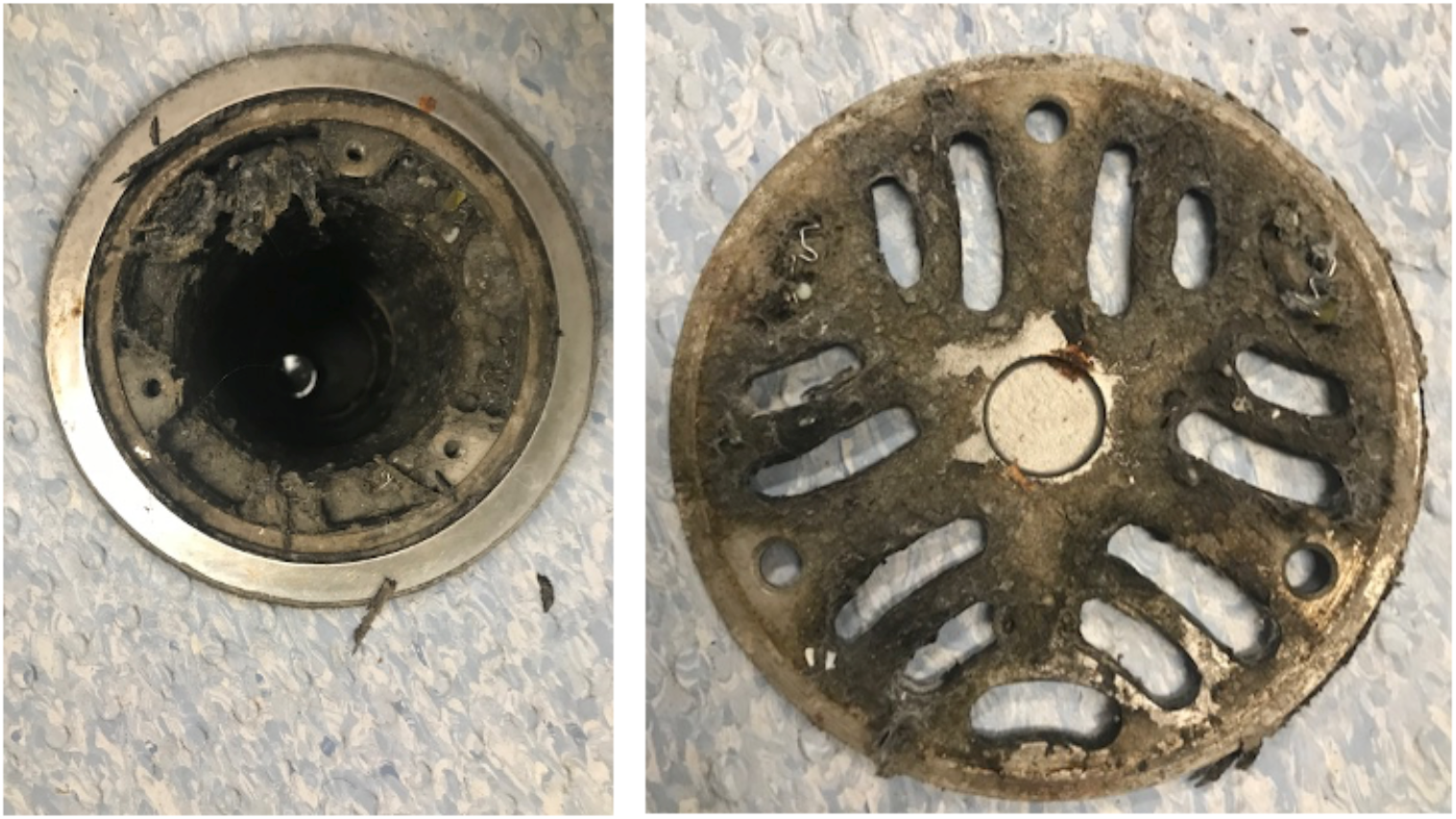
Burns bath 3 floor trap. an example of the biomass uncovered under the floor trap in a Burns Unit bathroom. Areas of high biomass (such as this one) were targeted for environmental screening.

An ST1050 CR-Ab was cultured using traditional methods from the environmental sample R5666, taken from a crack in a toilet seat being used by a patient colonized with the ST1050 CR-Ab. The depth of sequencing obtained from the same environmental sample, however, was not sensitive enough to be able to confidently detect the presence of the CR-Ab in the metagenomic data. The second positive ST1050 CR-Ab culture came from an environmental sample taken from an Antechamber room connected to patient rooms that had previously been colonized with ST1050 CR-Ab (R5864). Parallel metagenomic sequencing was also able to detect this same ST1050 CR-Ab in the environmental sample (Figure 5).

**Figure 5:**
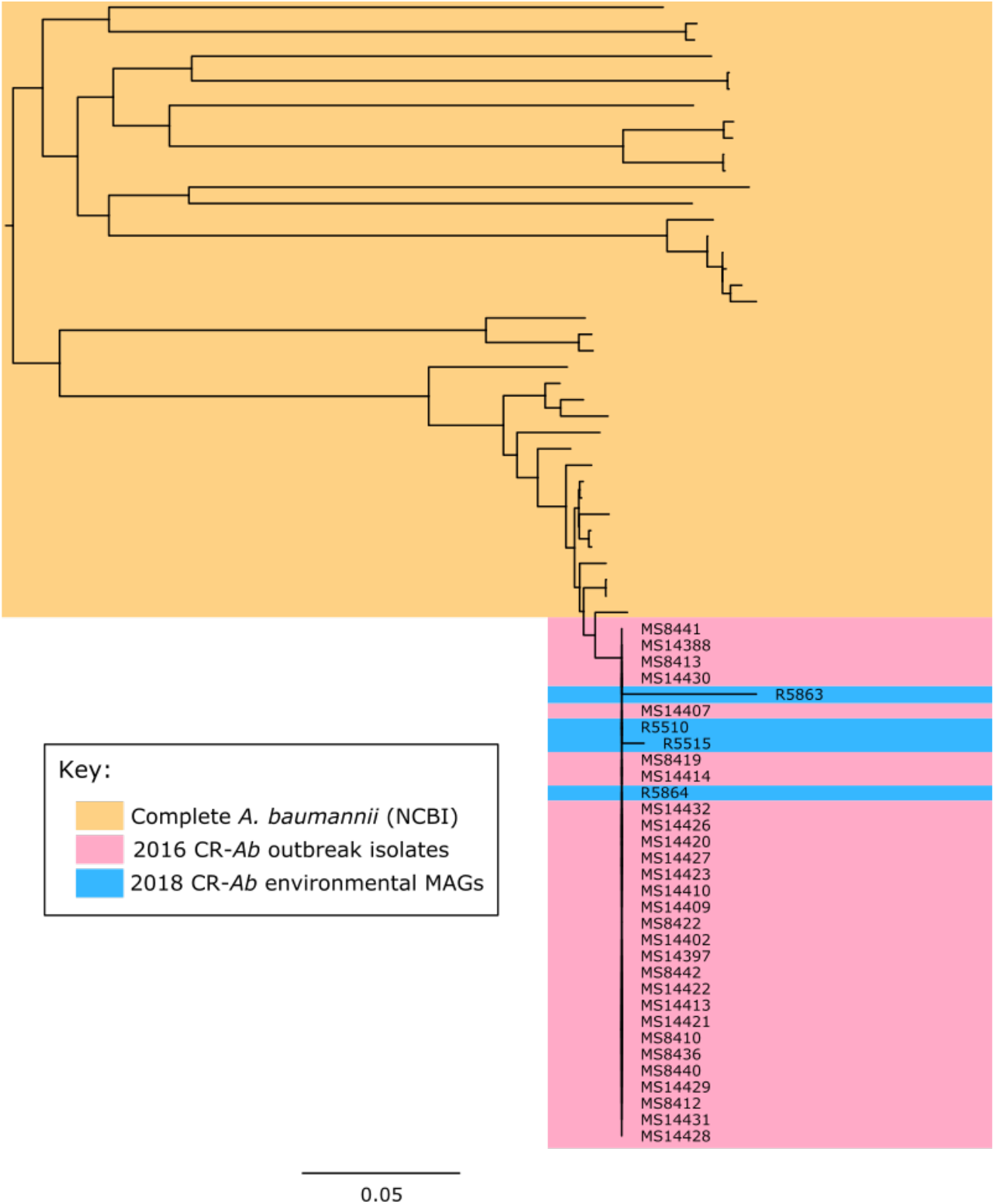
Clustering of MAGs with outbreak strains. Mid-point rooted core genome SNP phylogenetic tree contextualising the metagenome assembled genomes (MAGs) with *de novo* assemblies of the outbreak strains and publicly available complete *A. baumannii* genomes (yellow) showing clustering of the MAGs (blue) within the outbreak clade (pink).

Additionally, three other samples were found to have ST1050 CR-Ab based on metagenomic sequencing, despite being culture negative using traditional methods (figure 5, table S4). Samples R5515 (burns bath 2 floor trap water sample) and R5510 (burns bath 2 bath drain hole [interior]) were both positive for ST1050 CR-Ab. Both samples were taken at the same time from proximal locations, and patients colonized with ST1050 CR-Ab were using the burns bath in question. Samples R5863 was also positive for ST1050 CR-Ab, and was taken from the room previously occupied by a patient known to be colonized with ST1050 CR-Ab.

### Plumbing maintenance program implemented in response to genomic investigation

Shotgun metagenomic detection of the outbreak strain in the hospital plumbing provided the evidence base for implementation of a sustainable infection prevention strategy. Consequently, a routine plumbing maintenance program was instituted. Every month, pipes were soaked for 30-minute in sodium hydroxide, with additional soaking and scrubbing of drain plates. Since the implementation of these measures, no further cases of CR-Ab have been detected in the Burns Unit or Intensive Care Unit (ICU) following 28^th^ September 2018. Between December 2018-May 2019, three additional ST1050 cases were detected outside the ICU and Burns Unit and linked to the outbreak via routine surveillance under the Queensland Genomics Healthcare-associated Infections project. One case produced sputum and blood isolates from the same patient while the other two were detected at two different hospitals in South-East Queensland. Periodic environmental surveillance of CR-Ab in drains and plumbing in the Burns unit has been ongoing as of May 2020. No further detection of the outbreak strain has occurred since May 2019 (fig. S12).

### Significant reduction of risk following interventions

A total of 32 CR-Ab cases were recorded over 28 months in the pre-intervention period, compared to 4 CR-Ab cases over 21 months in the post-intervention period. All cases identified at pre-intervention period were admitted to the Burns or Intensive Care Units during their hospitalisation. Conversely, 3 out of the 4 CR-Ab cases detected in the post-intervention period had no obvious epidemiological link to exposure in the Burns and Intensive Care Units. The incidence rate post-intervention was 2 CR-Ab cases per year, significantly reduced from the pre-intervention incidence rate of 13 CR-Ab case per year (*p* < 0.001). The post-intervention CR-Ab incidence rate was reduced by 17% compared to the pre-intervention period (incidence rate reduction=0.17, 95% CI: 0.06-0.47).

### Discussion

CR-Ab are an increasingly dire threat to global public health. Their proficiency at surviving for long periods of time in environments whilst under antibiotic pressure is largely due to the positive selection of both intrinsic and acquired resistance and survival mechanisms. As such, they present a significant problem in health-care settings, which typically have high antibiotic use as well as a large cohort of vulnerable patients. Understanding the mechanisms behind their resistance and transmission, as well as their possible environmental reservoirs, is key to combating further colonization and infection in hospital settings. Here were present a comprehensive analysis of an outbreak of CR-Ab using isolate and environmental metagenomic sequencing to fully elucidate transmission, determine new cases rapidly and detect possible environmental reservoirs within the hospital.

Genomics is being rapidly established in clinical settings, particularly in response to outbreaks(*17, 18*). This is due not only to the higher discriminatory power that WGS provides, but also the complete picture that WGS captures by yielding the entire genome. The current cost and turnaround time for sequencing and analysis also make this type of investigation more feasible in nosocomial settings. In this study, initial sequencing of the outbreak CR-Ab isolates (and associated bacterial species) confirmed an already suspected outbreak, and so despite providing more insight into possible transmission routes, it did not greatly affect the infection control response. However, genomics superseded traditional methods when it came to [i] contextualizing outbreak isolates with previous CR-Ab strains from the hospital (to determine the likely source), and [ii] contextualizing new CR-Ab isolates as they appeared after the initial outbreak to determine whether there was an ongoing problem in the hospital. While having a slightly faster turnaround time, traditional methods alone would not have been able to confidently assess either of these scenarios. Regular meetings and reporting of the genomic results provided the hospital with actionable information and greater insight into the ongoing outbreak. These cross-disciplinary discussions facilitated the communication of complex genomic data into the clinical setting, providing guiding principles for subsequent WGS reporting of multidrug resistant bacterial pathogens at this hospital, and prompting the development of an interactive online visualization for communicating genomic epidemiology data (see movie S1).

In addition to providing evidence for related isolates, WGS was also a valuable tool for discerning unrelated isolates, in many cases preventing ward or operating theatre closures and mitigating the associated financial costs to the hospital(*19, 20*). It is plausible that with continued, ongoing sequencing of clinically significant bacteria in high-risk environments (e.g. ICU and Burns Unit) the risk of outbreaks could be reduced if evidence of transmission was detected early. During this study, we were able to detect transmission of an *E. hormaechei* unrelated to the outbreak at hand, but linked to a *bla*_IMP-4_ carbapenemase-producing Enterobacteriaceae (CPE) outbreak from the same hospital the year prior(*15*). We were also able to identify transmission of an unrelated meropenem-resistant *P. aeruginosa* isolate, highlighting how WGS can detect transmission well before it becomes known to staff. The discovery of four ST1050 outbreak isolates in late 2018 to early 2019 also highlights the importance of early detection and intervention to limit and control spread. These three patients had no exposure to the ICU or Burns Unit at any time during this outbreak, so we can only propose that the environment (i.e. the plumbing) and/or undetected transmission facilitated by other patients or healthcare workers (between the two additional hospitals) as the most likely source. Remarkably, as this manuscript was in preparation, we also discovered the same ST1050 outbreak strain from a patient readmitted to RBWH 18 months after positive blood and sputum cultures. Long-term carriage of *A. baumannii* has been observed previously(*21*). This finding not only implicates discharged patients in the spread between different hospitals but highlights their potential for reintroduction of the outbreak strain into settings where it has been previously eradicated.

Routine WGS can also lead to a reduction in the costs associated with responding to an established outbreak. A study of a similar outbreak in Brisbane determined the cost per patient related to the outbreak to be six-times higher than unrelated patients(*22*). However, the feasibility (i.e. access to sequencing facilities and analysis) of routinely sequencing multidrug-resistant organisms is not yet achievable for many hospitals.

Determining relatedness and transmission using genomics has historically relied on the number of core SNP differences between isolates(*23*-*25*). However, this approach has several flaws, including a general lack of consensus on SNP cutoffs and what number defines a related isolate within a particular species, as well as the fact that it largely ignores other genomic differences, such as large insertions, inversion and rearrangements. It also does not account for hypermutators, which we observed in the case of the *K. pneumoniae* isolate MS14418 (see supplementary methods and results 1). More recent methods have explored the use of transmission probabilities by taking into account isolation time and species mutation rate(*26*), but these methods appear more suited to outbreaks spanning large timeframes. Most studies to date that have used SNP distances have used them retrospectively and under research conditions, thereby avoiding the necessity to conform to standardized metrics and allow case-by-case judgments to be made on isolates. Moving forward, translating this approach into standardized clinical settings will likely present several hurdles. In our study, with the exception of the hypermutator strain MS14418 there was no ambiguity using SNP distances to determine relatedness due to the observed low mutation rate. However, because of this, many isolates were unable to be discriminated, with several identical at the core genome level. We were surprised that the initial polymicrobial nature of this outbreak enabled deduction of transmission routes by examining SNP differences between their respective companion *K. pneumoniae* isolates which appeared to have coinfected with the CR-Ab. However, all of these transmissions were from the index patient and were already recognized by the clinical team. In contrast, the spread of CR-Ab between the ICU and Burns Units in July could be traced to transmission of CR-Ab carrying a discriminatory SNP from the index patient to patient 10 in the Burns Unit with subsequent transmission of CR-Ab to Patient 11, 14 and 17 in the Burns Unit and Patient 13 in the ICU (Figure 1). Further work into identifying both SNPs and pan-genome markers (such as gain/loss of regions or movement of mobile elements) could assist in further characterizing this outbreak and others.

Metagenomic sequencing of the environment was able to identify several areas positive for ST1050 CR-Ab. In one case, metagenomic sequencing analysis and traditional culture methods were concordant and both identified the ST1050 CR-Ab. In all other cases, either traditional culture or metagenomic sequencing was able to recover the ST1050 CR-Ab, highlighting the advantage of using both methods during an outbreak. While metagenomic sequencing was able to recover more positive results than the traditional methods, it has several limitations, including the necessity for high bacterial loads (such that there is sufficient starting DNA to sequence) and the increased costs (in our study, we observed that at least 5 Gigabase pairs of sequencing data is required to get a basic amount of depth and sensitivity when looking for specific strains). In future, initial PCR from the environmental DNA targeting a known marker in the outbreak strain could help narrow the candidates for complete metagenomic sequencing. Further work is required to refine these methods and determine an accurate guideline, particularly as it relates to sequencing depth and sensitivity.

All of the positive sequencing and culture results from the environmental sampling were from areas presently or previously being used by patients colonized with the ST1050 CR-Ab. As such, we cannot be sure that the identified ST1050 CR-Ab was present in these environments prior to colonization, or if it was shed from the patient. Subsequent environmental sampling was carried out after each round of cleaning, and no CR-Ab was detected afterwards. It is most likely that the CR-Ab detected in the environmental reservoirs were shed from the patients, however this result does indicate the ease of transmission of this organism from colonized patients to fomites within the hospital, where they then might transmit to other areas or to hospital staff(*27*).

By using WGS to assist in a large outbreak of CR-Ab (and other MDR gram-negative bacilli) we show how genomics can be used to improve rapid respond measures and outbreak management, as well as provide in-depth characterization of the outbreak strains to establish a historical database that can be used to guide responses to future outbreaks. We also show how direct sequencing of environmental samples was able to detect evidence of the outbreak strain leading to key changes in infection control policy.

## Materials and Methods

### Study setting and patient inclusion

Primary isolates were obtained from patients admitted to the Royal Brisbane & Women’s Hospital (RBWH), a tertiary referral hospital with 929 beds in South-East Queensland, Australia. The RBWH has a 36 bed ICU providing highly specialist burns care for all of Queensland. The incidence of CR-Ab is low in Australian hospitals(*28*). All new CR-Ab strains are routinely stored in the clinical laboratory for future reference. For the outbreak investigation, any patient admitted to the RBWH who cultured CR-Ab from any clinical or screening specimen from May to August 2016 was identified as a case and included in the primary outbreak analysis. Any CR-Ab cases during the outbreak period were also included to determine if plasmid-mediated resistance and dissemination was relevant, with any MDR Gram-negative bacilli (including ESBL-producing *K. pneumoniae*, carbapenem-resistant *S. marcescens* or carbapenem-resistant *P. aeruginosa*) prospectively collected for further genomic analysis. Overall these included 28 CR-Ab, 3 carbapenem-sensitive *A. baumannii*, 10 *K. pneumoniae*, 7 *P. aeruginosa*, 4 *S. marcescens* and 3 *Enterobacter cloacae* (the *E. cloacae* were isolated in relation to a previous outbreak in the same hospital(*15*)). Stored CR-Ab isolates from a previous outbreak in 2006(*6*), as well as other sporadic cases imported from overseas to the RBWH during 2015/2016 (prior to the outbreak) were included for further analysis. These included 17 historical CR-Ab isolates from earlier in 2016 (n=3), 2015 (n=2) and between 2000-2006 (n=12). *A. baumannii* identified from the outbreak until mid-2018 were also included in the analysis during continued surveillance and infection control monitoring. These included 3 carbapenem-sensitive *A. baumannii* and 19 CR-Ab isolates. A complete list of all isolates is provided in supplementary data 1.

### Antimicrobial susceptibility testing

All bacterial isolates were identified by matrix-assisted laser desorption/ionization mass spectrometry (MALDI-TOF) (Vitek MS; bioMérieux, France). Antimicrobial susceptibility testing was carried out using Vitek 2 automated AST-N426 card (bioMérieux). For the first 8 sequential CR-Ab isolates, additional susceptibility testing was undertaken using Etest to determine MICs for meropenem, imipenem, colistin, tigecycline, fosfomycin, amikacin, sulbactam, doxycyline and ceftolozane/tazobactam, with disk diffusion to determine susceptibility to aztreonam and ceftazidime/avibactam. Carbapenemase activity was assessed by the use of the Carba-NP test (RAPIDEC; bioMérieux) and screened for the presence of common carbapenemases found in Enterobacteriaceae using an in-house multiplex real-time PCR (that targets NDM, IMP-4-like, KPC, VIM and OXA-48-like carbapenemases). Once it became clear that all the outbreak strains had an identical antibiogram, susceptibility testing was confined to the Vitek 2 automated AST-N426 panel with MICs to tigecycline, doxycycline and colistin determined by Etest (as the only susceptible agents).

### Bacterial culturing and genomic DNA extraction

All isolates were grown on horse blood agar at 37°C overnight. For all historical and outbreak isolates collected between May-September of 2016, colonies were scraped from plates and resuspended in 5 mL Luria Bertani (LB) broth. 1.8 mL of resuspension was use for DNA extraction using the UltraClean^®^ Microbial DNA Isolation Kit (MO BIO Laboratories) as per manufacturer’s instructions. All isolates collected after September 2016 were extracted using the DSP DNA Mini Kit on the QIAsymphony SP (Qiagen).

### Isolate whole genome sequencing (WGS)

Illumina WGS of suspected outbreak patient isolates and historical CR-Ab isolates was performed in four batches of between 10 and 18 samples between June and August 2016 at the Australian Centre for Ecogenomics (ACE), The University of Queensland (see supplementary methods and results 1). One CR-Ab isolate (MS14413) and one *K. pneumoniae* isolate (MS14393) were selected for sequencing with Pacific Biosciences (PacBio) Single Molecule Real-Time (SMRT) sequencing on an RSII machine (see supplementary methods and results 1). Subsequent Illumina WGS was carried out at Queensland Forensic Scientific Services (QFSS) (see supplementary methods and results 1).

### Quality control and assembly of WGS data

Illumina raw reads were checked for contamination using Kraken(*29*) v0.10.5-beta and quality using FastQC v0.11.5 (www.bioinformatics.babraham.ac.uk/projects/). Raw reads were filtered for reads less than 80 bp and quality score less than 5 using Nesoni clip v0.130 (https://github.com/Victorian-Bioinformatics-Consortium/nesoni). Some reads required further hard trimming with Nesoni clip (10 bp from start, 40 bp from end). Isolates were assembled using SPAdes(30) v3.6.0 at default settings. Contigs less than 10x coverage were removed using a custom script. Assembly metrics were checked for quality using Quast(*31*) v4.3 (see supplementary data 1). Details of the PacBio genome assembly and annotation can be found in the supplementary methods (supplementary methods and results 1).

### Genomic analysis and clinical reporting

Between June and August 2016, four reports of detailed bioinformatic analyses were prepared in response to available Illumina data for *A. baumannii, K. pneumoniae, P. aeruginosa, S. marcescens* and *Enterobacter cloacae* patient isolates. Comparative genome analysis using variant calling, phylogenetic reconstruction, transmission pathway prediction, multilocus sequence typing (MLST) resistance gene prediction and plasmid characterization used in the clinical reports are given in supplementary methods (see supplementary methods and results 1). For subsequent analyses of the final genome dataset updated or alternative software was used as described below.

Core single nucleotide polymorphisms (SNPs) were identified using Snippy(32) (v4.3.6) at default settings and trimmed reads against the complete chromosomes for MS14413 (CR-Ab) and MS14393 (*K. pneumoniae*). Parsnp (v1.2) (at default with “-c” flag) was used to visualize phylogenetic relatedness between the outbreak CR-Ab and the historical *A. baumannii* isolates. Multilocus sequence typing (MLST) was performed using mlst(*33*) v2.6 (https://github.com/tseemann/mlst) against the draft assemblies. The Oxford MLST scheme was used for the CR-Ab isolates(*34*). Resistance genes were identified using Abricate(*35*) v0.6 against the ResFinder database(*36*) (accessed August 18^th^, 2017). Abricate was also used to determine plasmid types using the PlasmidFinder database(*37*) (accessed August 18^th^, 2017). Comparative analyses were completed using the Artemis Genome browser and the Artemic Comparison Tool (ACT). Figures were constructed using EasyFig(*38*), BRIG(*39*) and FigTree(*40*).

### Metagenomic sequencing and analysis

Metagenomic sequencing of environmental samples and analysis was conducted as described previously(*15*). Briefly, swab and water samples from the ICU and Burns Unit were collected in July 2018. DNA was extracted using the Qiagen DNeasy Powersoil extraction kit and sequenced at the Australian Centre for Ecogenomics on an Illumina NextSeq500.

All samples were screened for species using Kraken(*29*) v1.0 and resistance genes using SRST2(*41*) v0.2.0 against the ARG-ANNOT(*42*) database. Mash(*43*) v1.1.1 was used at default settings to screen Illumina reads for each samples against our reference CR-Ab sketch (MS14413). Samples that shared ≥90% of hashes were mapped to the reference sequence. Mapped reads were parsed and *de novo* assembled using SPAdes(*30*) v3.11.1 for MLST analysis using mlst(*33*) v2.16.2 and nucleotide comparison using ACT(*44*) and BRIG(*39*).

### Risk reduction assessment

We aimed to estimate the reduced risk of patient colonization following the identification of ST1050 CR-Ab by environmental metagenomic sequencing and the initiation of enhanced decontamination of hospital plumbing. The incidence rate of CR-Ab was measured pre-intervention and post-intervention. The point of intervention was defined as the targeted initiation of routine plumbing maintenance program within the Burns and Intensive Care units in August 2018. The intervention was expected to generate immediate results with no lag time. The pre-intervention period was defined as May 2016 to August 2018 and post-intervention period as September 2018 to May 2020. All CR-Ab cases recorded in the hospital during these periods were included. Patients admitted to the Burns and Intensive Care units underwent standard clinical swabbing for surveillance and laboratory method for testing did not change over the study period. Statistical analyses were performed on Rv3.5.1.

## Supplementary Materials

### Supplementary methods and results 1

(supplementary_methods_and_results_1.pdf): *Supplementary results and methods:* additional methods and supplementary results to the main text.

Supplementary data 1 (supplementary_data_1.xlsx): *Strain list:* A list of all strains analysed in this study, with their accessions, assembly metrics and associated metadata. Supplementary report 1 (supplementary_report_1.pdf): *First genomic report:* original genomic report on first batch of WGS data delivered June 22, 2016 (redacted for privacy).

Supplementary report 2 (supplementary_report_2.pdf): *Fourth genomic report:* original clinical report on fourth batch of WGS data delivered August 29, 2016 (redacted for privacy), annotated to highlight alterations to report design in consultation with clinical staff.

Movie S1 (supplementary_movie_1.mp4): *Communication of outbreak with HAIviz:* video of outbreak progression produced using Healthcare-Associated Infection visualization tool (http//:HAIviz.beatsonlab.com), an interactive web-based visualization tool designed to communicate infectious disease genomic data from local outbreaks to healthcare professionals. The dashboard shows outbreak timeline, a local map, patient locations, predicted transmission links and the genetic relationships of isolates based on WGS data.

## One sentence summary

By using prospective whole genome sequencing (WGS) combined with detailed reporting and environmental metagenomic sequencing, we were able to fully characterize a polymicrobial outbreak in critical care by determining the main causative outbreak strain (an ST1050 carbapenem-resistant *Acinetobacter baumannii)* and identifying key reservoirs in the environment to resolve the outbreak.

## Data Availability

The datasets supporting the conclusions of this article are available in the short read archive (SRA) repository, under the following Bioprojects: the complete genomes for MS14413 (GenBank: CP054302.1) and MS14393 (GenBank: CP054303-CP054305) have been deposited under the Bioprojects PRJNA631347 and PRJNA631348, respectively. All isolate Illumina sequencing reads have been deposited under the Bioproject PRJNA631491. All metagenomic Illumina sequencing reads have been deposited under the Bioproject PRJNA631351.

## Declarations

### Ethics approval and consent to participate

Ethics approval was provided by the RBWH HREC as a low-risk study with waiver of consent (HREC/16/QRBW/581).

### Consent for pub/ication

Not applicable.

### Availability of data and materials

The datasets supporting the conclusions of this article are available in the short read archive (SRA) repository, under the following Bioprojects: the complete genomes for MS14413 (GenBank: CP054302.1) and MS14393 (GenBank: CP054303-CP054305) have been deposited under the Bioprojects PRJNA631347 and PRJNA631348, respectively. All isolate Illumina sequencing reads have been deposited under the Bioproject PRJNA631491. All metagenomic Illumina sequencing reads have been deposited under the Bioproj ect PRJNA631351.

### Competing interests

PNAH has received research grants from MSD, Sandoz and Shionogi Ltd, outside of the submitted work, and speaker’s fees from Pfizer paid to The University of Queensland. DLP reports receiving grants and personal fees from Shionogi and Merck Sharp and Dohme and personal fees from Pfizer, Achaogen, AstraZeneca, Leo Pharmaceuticals, Bayer, GlaxoSmithKline, Cubist, Venatorx, and Accelerate. JL has received personal fees from Pfizer and MSD and grants from MSD paid to The University of Queensland. The other authors have no conflicts of interest to declare.

### Funding

LWR was supported by an Australian Government Research Training Program (RTP) Scholarship. SAB, PNAH and MAS were supported by fellowships from the Australian National Health and Medical Research Council (GNT1090456, GNT1157530 and GNT1106930, respectively). The work was supported by VC Strategic Intitiative Funding from The University of Queensland (2016-2018) and grants from the Queensland Genomics Health Alliance (now Queensland Genomics), Queensland Health, Queensland Government (https://queenslandgenomics.org/projects/round-2/infectious-disease-portfolio/#healthcare-associated-infections-project). The funders had no role in study design, data collection and interpretation, or the decision to submit the work for publication.

### Authors’ contributions

LWR, PNAH and SAB designed study. PNAH, GRN, NG, JM and JL coordinated patient inclusion and isolate collection from the hospital. LWR and TH collected environmental samples. TH and KH coordinated infection control response. LWR, BMF and WL performed experiments. LWR, SAB, BMF and PNAH prepared clinical reports. All authors contributed to the interpretation of results. SAB, PNAH, MAS and DP supervised aspects of the project and provided essential expert analysis. LWR, PNAH, SAB, WL, and TH wrote the manuscript. All authors read and approved the final manuscript.

## Acknowledgements

We acknowledge all staff at the Royal Brisbane and Women’s Hospital and Pathology Queensland who were involved in patient care and the clinical response to the outbreak described in this study. We acknowledge the facilities, and the scientific and technical assistance of staff at the Australian Centre for Ecogenomics Sequencing Facility (The University of Queensland (UQ)) and the Public Health Microbiology Laboratory at Queensland Forensic and Scientific Services (Queensland Health). We thank Thomas Cuddihy (QFAB Bioinformatics and Research Computing Centre, UQ) for high-performance computing support. We thank Kate Peters (Schembri lab, UQ) For technical assistance preparing genomic DNA.

## Supplementary Material

### Supplementary Methods

#### Illumina sequencing

All isolates collected between May-September 2016 (and all historically collected isolates) were sequenced at the Australian Centre for Ecogenomics (ACE) Sequencing Service, University of Queensland, Brisbane, Australia. DNA was quantitated using Qubit and libraries prepared using Nextera XT library prep (Illumina) with Nextera XT/V2 Indexes, as per manufacturer’s instructions. Resulting libraries were quantitated with either qPCR or Tapestation, pooled and each sample loaded onto either 1/100^th^ or 1/200^th^ of a flow cell and sequenced on the NextSeq (Illumina) using a 2 x 150bp High Output V2 kit.

All subsequent isolates (from October 2016 onwards) were sequenced at the Public Health Microbiology Laboratory at Queensland Forensic and Scientific Services, Brisbane, Australia. All libraries were prepared using the Nextera XT DNA preparation kit (Illumina) and sequencing was performed on a NextSeq 500 (Illumina) with 2x150bp chemistry, NextSeq Midoutput kit v2.5.

#### Bioinformatic analysis for clinical reporting

Methodologies for bioinformatic analysis and communication of Illumina WGS data during primary outbreak in 2016 (June 22, July 16, Aug 2, Aug 29) are outlined below: Quality control and *de novo* assembly of Illumina WGS data and comparative genome analysis were carried out as described in the main document. Raw reads were analysed using Nullarbor (https://github.com/tseemann/nullarbor) to determine MLST, antibiotic resistance gene profile, and core SNP phylogeny using species-specific reference sequences. Closest publicly available complete genomes were chosen as reference sequences where available (*Acinetobacter baumannii* Global Clone (GC) 2 strain 1656-2 (GI:384129960); *Klebsiella pneumoniae subsp. pneumoniae* MGH 78578 (GI:150953431); *Serratia marcescens* WW4 (GI:448239774); *Pseudomonas aeruginosa* PA1R (GI:558665962). For *Enterobacter cloacae* the reference was the concatenated draft genome of *Enterobacter cloacae* Ecl1 (GenBank: JRFQ01000000; now reassigned as *E. hormaechei*), an ST90 strain isolated from a burns patient at the RBWH ICU in 2015. Antibiotic resistance gene content and MLST was further checked using srst2(*41*) against the ARG-ANNOT(*42*) database and the Oxford MLST scheme(*34*), respectively. Plasmid Inc Typing was done based on the relaxase gene as described by Compain *et al*. (*45*).

SNP differences between strains were determined using Nesoni (https://github.com/Victorian-Bioinformatics-Consortium/nesoni) and evolutionary relationships were determined shown as phylograms or Eburst-style matrices in which nodes of identical isolates were separated by branches representing one or more core SNP differences radiating from a founder (index) node. This format provided consistency across reports, enabling a progressive expansion of the display figure from a common anchor as each WGS batch was reported, in contrast to phylograms where topology and isolate order could change substantially as the data was updated. For example Supplementary Figure 2 is a close approximation of the CR-Ab tree reported in August 2016 and forms the anchor to Figure 3B which shows all ST1050 CR-Ab in the study.

Methodologies used for subsequent reports (Nov 4 2016, Mar 9 2017, Jun 20 2017, Oct 10 2017) were essentially the same except that the concatenated draft genome of ST1050 CR-Ab MS8436 or MS14413 was used as reference sequences, Abricate (v0.6) with ResFinder was used for antibiotic gene prediction, and from 2017 Nesoni implemented Bowtie for alignment instead of SHRiMP.

All 2018 CR-Ab isolate genomes were initially reported to RBWH as part of an Infectious Diseases demonstration project in WGS surveillance of MDR bacteria in hospitals (encompassing most of this authorship group) funded by the Queensland Genomics Health Alliance (now Queensland Genomics), Queensland State Government, Australia.

#### Pacific Biosciences sequencing

One CR-Ab isolate (MS14413) and one *K. pneumoniae* isolate (MS14393) were selected for sequencing with Pacific Biosciences (PacBio) Single Molecule Real-Time (SMRT) sequencing. Isolates were grown on LB agar at 37°C overnight. A single colony was used to inoculate 10 ml LB broth, grown overnight at 37°C (shaking 250 rpm). DNA was extracted using the UltraClean^®^ Microbial DNA Isolation Kit (MO BIO Laboratories) as per manufacturer’s instructions. 20kb SMRTbell libraries were prepared using P6 polymerase and C4 sequencing chemistry with 7kb size selection with BluePippin. Final polymerase bound libraries were sequenced using 1 SMRT cell each on a PacBio RSII instrument at the University of Queensland Centre for Clinical Genomics, Translational Research Institute, Brisbane, Australia.

#### Pacific Biosciences genome assembly and annotation

PacBio genomes were assembled using Canu(*46*) v1.3 and manually closed using Artemis(*47*). A large duplicated region of ~40 kb was identified in the CR-Ab isolate and resolved using read-mapping and PCR at unique borders of the duplication (see below). The SMRT Analysis suite (v7.0.1.66975) was used to generate methylated motif summaries and polished assemblies using the PacBio reads. Indels were further corrected with Pilon(*48*) v1.22 using the trimmed Illumina reads mapped to the assembly using BWA(*49*) v0.7.16a-r1181. Complete PacBio genomes were annotated using Prokka(*50*) v1.12-beta. Insertion sequences were identified using ISSaga(*51*). Frameshifts were corrected by manually inspecting read pileup at suspected positions identified using NCBI microbial genome submission check (https://www.ncbi.nlm.nih.gov/genomes/frameshifts/frameshifts.cgi).

#### Closing the genome of CR-Ab strain MS14413

*De novo* assembly of the CR-Ab strain MS14413 PacBio reads using Canu resulted in two contigs: one large contig (~4 Mb) and one small contig (~65 kb). The large contig corresponded to the CR-Ab chromosome and could be circularized. Comparison of both contigs to each other using the Artemis Comparison Tool (ACT) determined that the smaller contig matched the ends of the chromosome perfectly except for a duplicated ~4.8 kb region. Rearrangement of this region onto the contig ends and re-mapping of both PacBio and Illumina reads to the chromosome (using BWA and blasr, respectively) resolved the ~4.8 kb region but simultaneously identified approximately twice as much read depth across a ~40 kb region on the chromosome, indicative of a large collapsed repeat in our initial assembly. Comparative analysis of the smaller contig (using ACT) followed by PCR at unique borders of the suspected tandem duplication (fig. S1, Table S1) enabled resolution of the region as two copies of ~41 kb. Further mapping with both the Illumina and PacBio reads (using BWA(*49*) and blasr(*52*), respectively) confirmed the tandem duplication, which has been included in the complete genome of CR-Ab strain MS14413 (GenBank: CP054302.1).

## Supplementary Results

### ~40 kb tandem duplication in CR-Ab genome

PacBio sequencing of CR-Ab isolate MS14413 identified a large tandem duplication of approximately ~40 kb, encompassing Tn2006 as well as part of the capsule (K) region (Figure 3, main text). Analysis of the other CR-Ab isolates using the Illumina *de novo* assemblies found evidence for this duplication in only two related colistin resistant isolates from the index patient (MS14413 [PacBio] and MS14402), suggesting that this duplication arose once and was maintained by a sub-population of CR-Ab within this patient for at least 36 days. The effect of this duplication on fitness is as of yet unknown.

### Transmission of *K. pneumoniae* parallel to *CR-Ab* transmission

Ten ESBL-producing *K. pneumoniae* isolates were collected from 5 patients during the outbreak and were all found to be ST515. Nine of the ten isolates differed by less than 10 core SNPs, indicating direct transmission within the ICU ward (fig. 8). A single isolate from the index patient (MS14418) was found to have an additional 61 core SNPs, consistent with a hypermutator phenotype. Indeed, further investigation of this isolate found an in-frame 9 bp deletion in *mutH*, resulting in the loss of 3 amino acids from this protein (fig. S9). The exact mechanism behind this deletion in *mutH* and its relationship to the hypermutator phenotype remains to be further elucidated.

All ESBL-positive *K. pneumoniae* isolates had identical antibiotic resistance gene profiles, including the ESBL gene bla_CTX-M-15_, other β-lactamases (*bla*_TEM_, *bla*_OXA-1_) and the aminoglycoside resistance gene *aac(6')Ib-cr*. Two isolates from the index patient (MS14393 and MS14418) developed resistance to carbapenems, which was likely due to an *IS10R* insertion in the outer membrane porin gene *ompK36* (fig. S8). Isolate MS14433 (from patient 16) also contained an IS*10R* inserted into *ompK36*, however the insertion was found to be close to the 5’ boundary of the *ompK36* gene and based on *in silico* analysis there was no evidence that it affected the function of the resulting protein. Isolate MS14393 (from the index patient) also possessed a nonsense mutation in the antibiotic resistance protein repressor gene *marR*, which could contribute to its overall resistance to antibiotics.

### The *K. pneumoniae* isolates carry two plasmids involved in antibiotic resistance and virulence

A single *K. pneumoniae* isolate from the index patient (MS14393) was sequenced using PacBio long-read sequencing to generate a high-quality reference genome, consisting of a 5,492,431 bp chromosome, a 216,803 bp IncF plasmid (pMS14393A), and a 125,232 bp IncA/C plasmid (pMS14393B). Most of the antibiotic resistance genes resided on the IncA/C plasmid in two main loci (fig. S6). The larger IncF plasmid did not contain any antibiotic resistance genes, but did harbor several heavy metal resistance operons, including resistance to copper, arsenic and mercury (fig. S10).

### Genomic factors affecting adhesion, biofilm formation and motility

Analysis of SNP differences between the ST1050 CR-Ab isolates found a single SNP resulting in a reversion of a nonsense mutation to a functional amino acid codon in the gene *filB* within the *fil* operon; a putative type 3 filamentous fimbriae. This reversion was obscured in the Snippy analysis, as one isolate (MS14422) was heterozygous at this position, resulting in it being masked from the core SNP analysis. This reversion corresponded to the latter half of the outbreak, including all CR-*Ab* isolates taken after the 4^th^ of July 2016 (with the exception of MS14413 [6/7/16], MS14438 [12/9/16] and SS17M5036 [17/5/17]) (fig. S4). As stated, MS14422 appeared to have both alleles at this position and represents either (i) a transitioning population reverting from a functional codon back to a stop codon, or (ii) a mixed population of both allele types. Due to the unusual nature of this nonsense mutation reversion, we downloaded all available complete publicly available *A. baumannii* strains from NCBI (accessed 15-11-2018) and inspected the *filB* region. This analysis showed that there were multiple *A. baumannii* strains with disrupted *filB* genes, caused not only by nonsense mutations, but also interruption by insertion sequences and frameshift mutations (fig. S4). Many of the publicly available strains also appeared to have functional *filB* genes, with reversion from a stop codon to a functional gene possible based on the phylogeny. While not much is known about this fimbriae in *A. baumannii*, several of the genes within this operon were shown to be down-regulated in community settings(*53*), suggesting that it may not be required in biofilms or stable bacterial populations. It is possible that it increases survivability or transmission throughout the environment, however further work is required to determine the phenotypic qualities of this mutation.

Conversely, and IS*Aba125* element was identified upstream of the *csu* operon in the complete genome of MS14413. The *csu* operon is a well-characterized chaperone-usher pili assembly system involved in biofilm formation(*54*). It is possible that this insertion sequence (IS) is driving enhanced expression of this operon, promoting adhesion to abiotic surfaces and encouraging biofilm formation. Unfortunately, due to the nature of the draft *de novo* assemblies derived from short-read sequencing, we were unable to fully characterise this insertion in all isolates. Further work is required to completely characterise the position of this IS in all outbreak isolates.

### No transmission of *P. aeruginosa* or *S. marcescens* from the index patient was observed

*P. aeruginosa* isolates from the index patient were found to be ST979 and all carried 5 antibiotic resistance genes (*aph(3’)-IIb, bla*_OXA-50_, *bla*_PAO_, *catB7* and *fosA*) (Supplementary Table 2). The final *P. aeruginosa* isolate from the index patient (MS14412) was found to be more resistant to carbapenems, likely due to a nonsense mutation in the outer membrane porin *oprD*, as well as a non-conservative amino acid change in the response regulator *parR*. Initial *S. marcescens* isolates appeared to only carry *aac(6’)-Ic, bla*_SRT-2_, *oqxB*, and *qnrE*. However, later acquisition of an IncHI2 plasmid in two *S. marcescens* isolates was associated with large number of additional resistance genes, including the ESBL *bla*_CTX-M-15_ (Supplementary Table 3).

**Supplementary Table 1:**
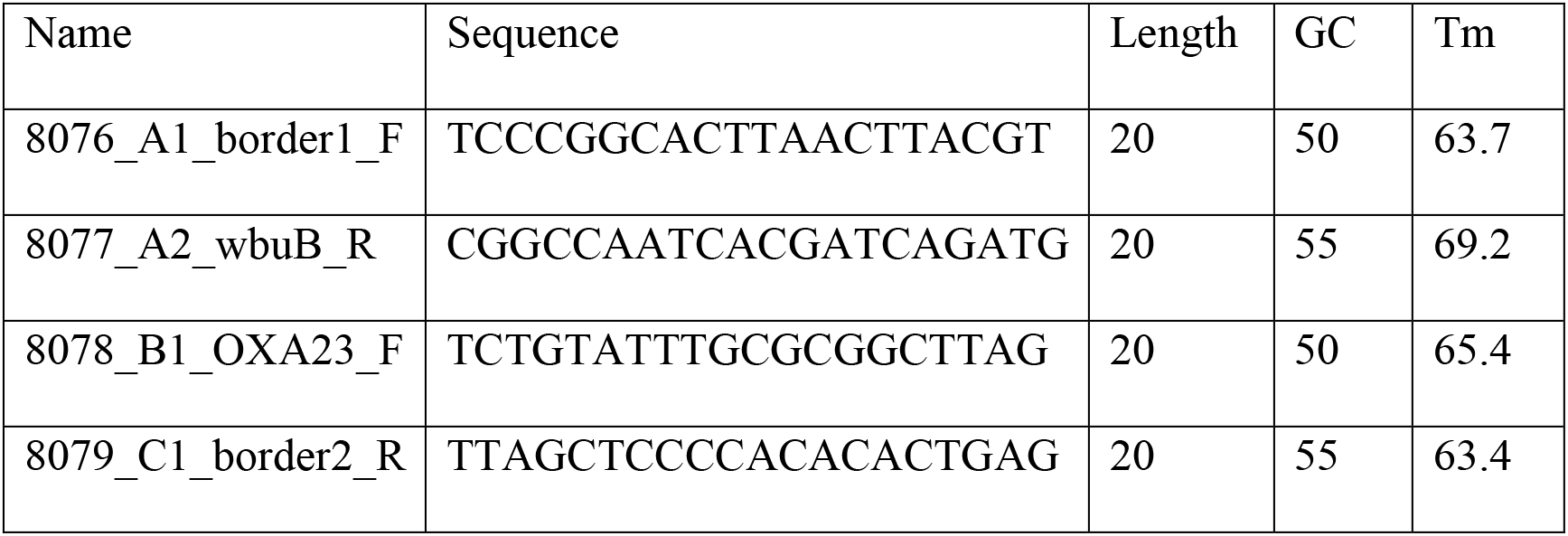
Primers used to resolve large duplication in CR-Ab isolate MS14413

**Supplementary Table 2:** Antibiotic resistance genes in *P. aeruginosa* isolates from the index patient: tick denotes presence

|  | aph(3')-IIb_1 | blaOXA-50_1 | blaPAO_4 | catB7_1 | fosA_1 |
| --- | --- | --- | --- | --- | --- |
| MS14395 | ✓ | ✓ | ✓ | ✓ | ✓ |
| MS14399 | ✓ | ✓ | ✓ | ✓ | ✓ |
| MS14403 | ✓ | ✓ | ✓ | ✓ | ✓ |
| MS14412 | ✓ | ✓ | ✓ | ✓ | ✓ |

**Supplementary Table 3:**
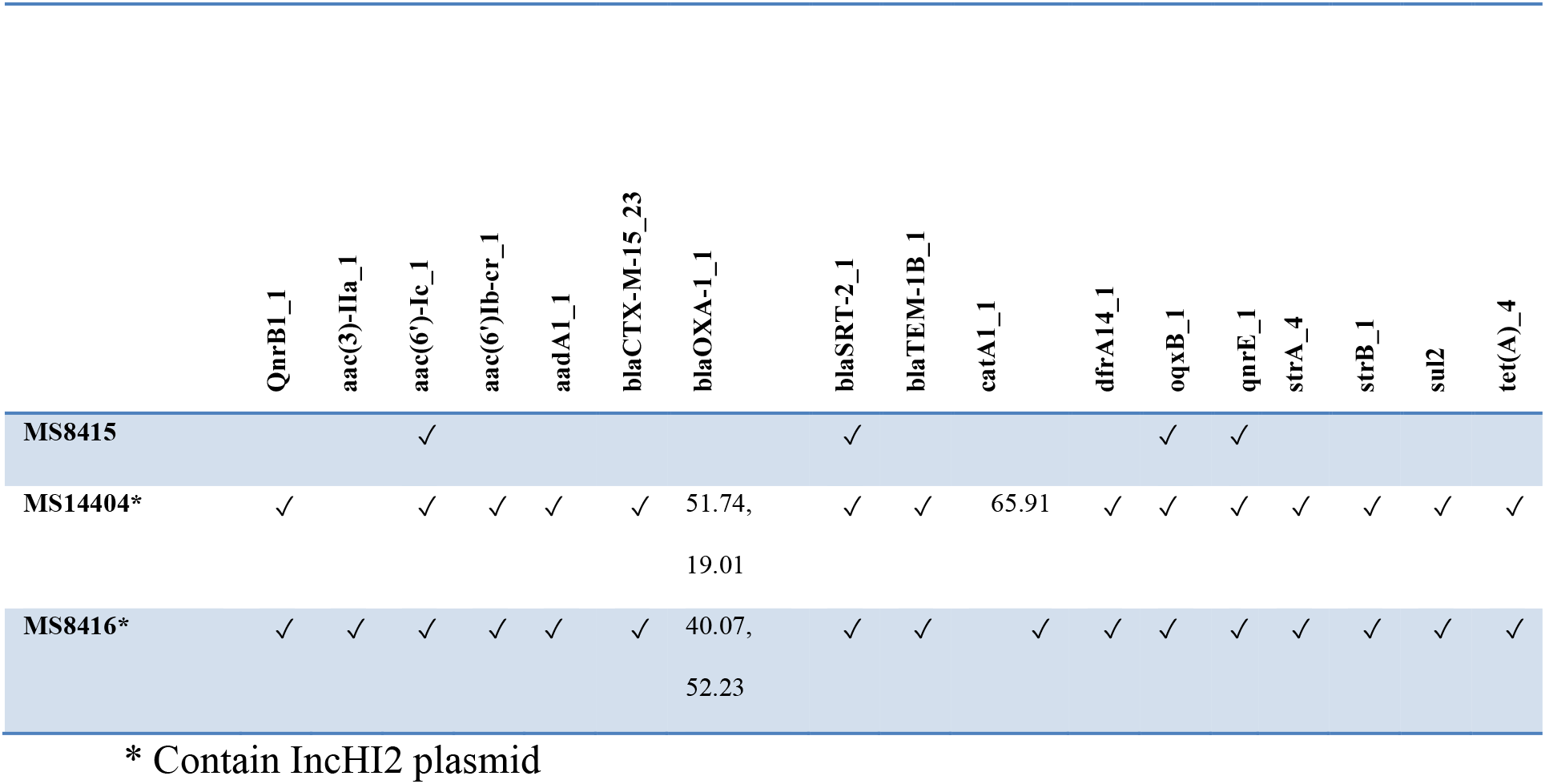
Antibiotic resistance genes in *S. marcescens* isolates from the index patient: tick denotes presence; number denotes %nucleotide coverage (where two numbers separated by a comma represents a split blast result)

**Supplementary Table 4:** Positive culture and sequencing results from 2018 environmental sampling.

| Sample | Isolation date | Culture result | Sequencing result | # reads total in sample | # reads mapped to MS14413 | % reads | # Contigs in *de novo* assembly |
| --- | --- | --- | --- | --- | --- | --- | --- |
| R5666 | 16/08/2018 | ST1050 CR-Ab | No CR-Ab | n/a | n/a | n/a | n/a |
| R5515 | 26/07/2018 | No CR-Ab | ST1050 CR-Ab | 43582216 | 444810 | 1.0% | 4173 |
| R5510 | 26/07/2018 | No CR-Ab | ST1050 CR-Ab | 51421258 | 3134890 | 6.1% | 8125 |
| R5863 | 02/10/2018 | No CR-Ab | ST1050 CR-Ab | 41402234 | 369031 | 0.9% | 10011 |
| R5864 | 02/10/2018 | ST1050 CR-Ab | ST1050 CR-Ab | 38635392 | 2634975 | 6.8% | 1326 |

**Supplementary Figure 1:**
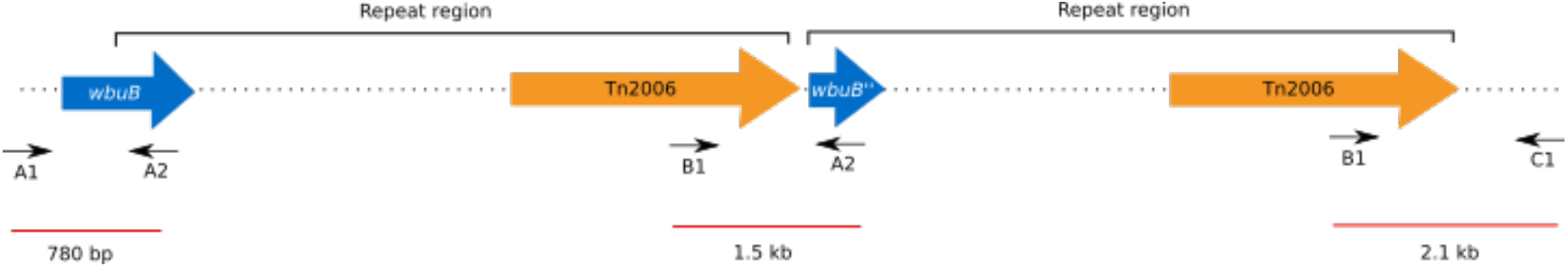
Primer binding regions to resolve large duplication in CR-Ab isolate MS14413 (not to scale)

**Supplementary Figure 2:**
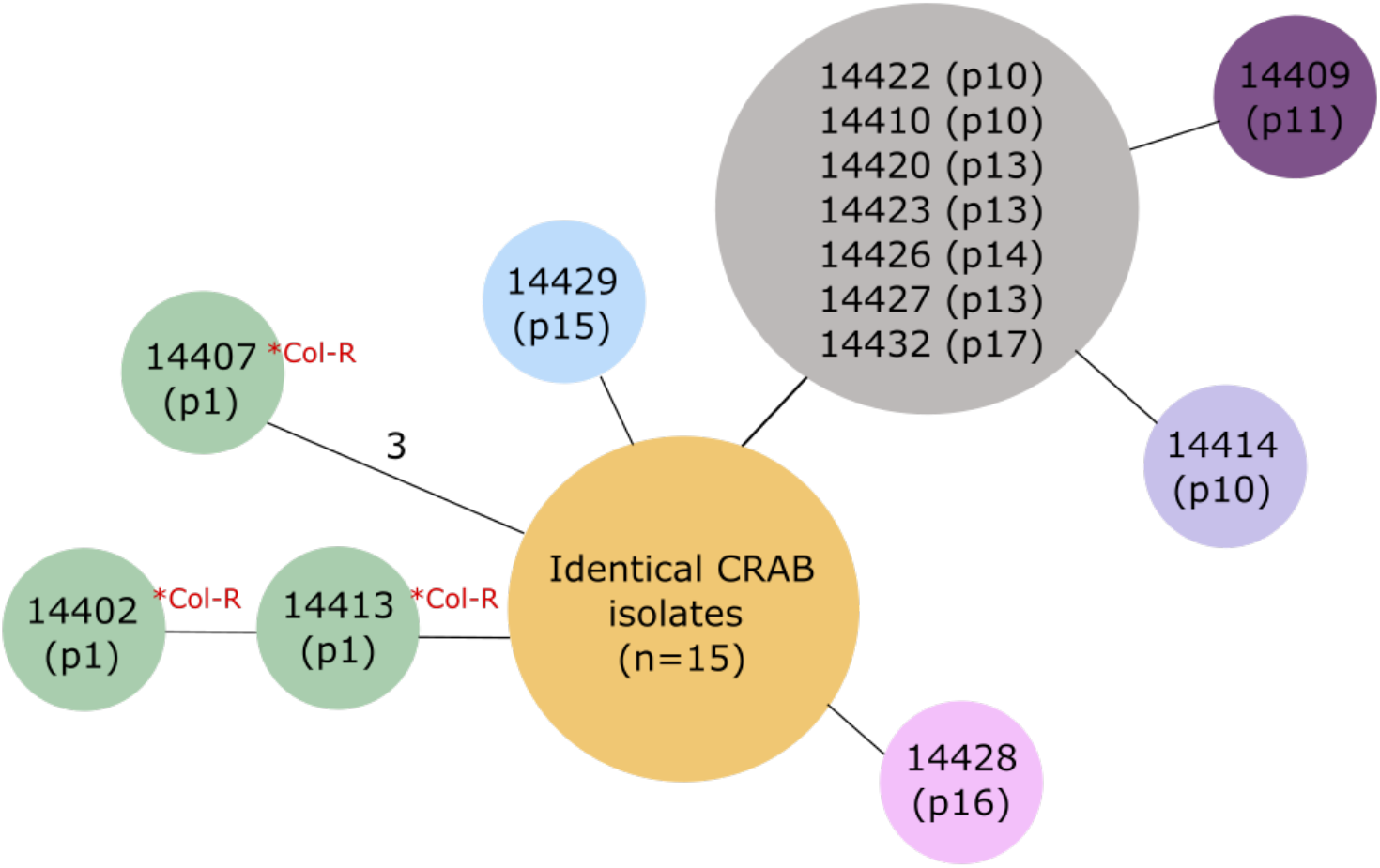
Relationship matrix of outbreak ST1050 CR-Ab isolates from 2016. isolates within the same circle are identical at the core SNP level. Black lines indicate 1 SNP difference, except where stated otherwise. Patient numbers denoted with “p” and the corresponding number. “Col-R” denotes isolates with colistin resistant SNPs. The yellow circle includes 15 isolates identical at the core SNP level from patients 1-9 and 12: MS14431, MS14430, MS14397 (p1); MS8410, MS8412 (p2); MS8422 (p3); MS8413 (p4); MS8419 (p5); MS8436, MS8440 (p6); MS8442 (p7) MS8441 (p8), MS14388 (p9) and MS14421 (p12).

**Supplementary Figure 3:**
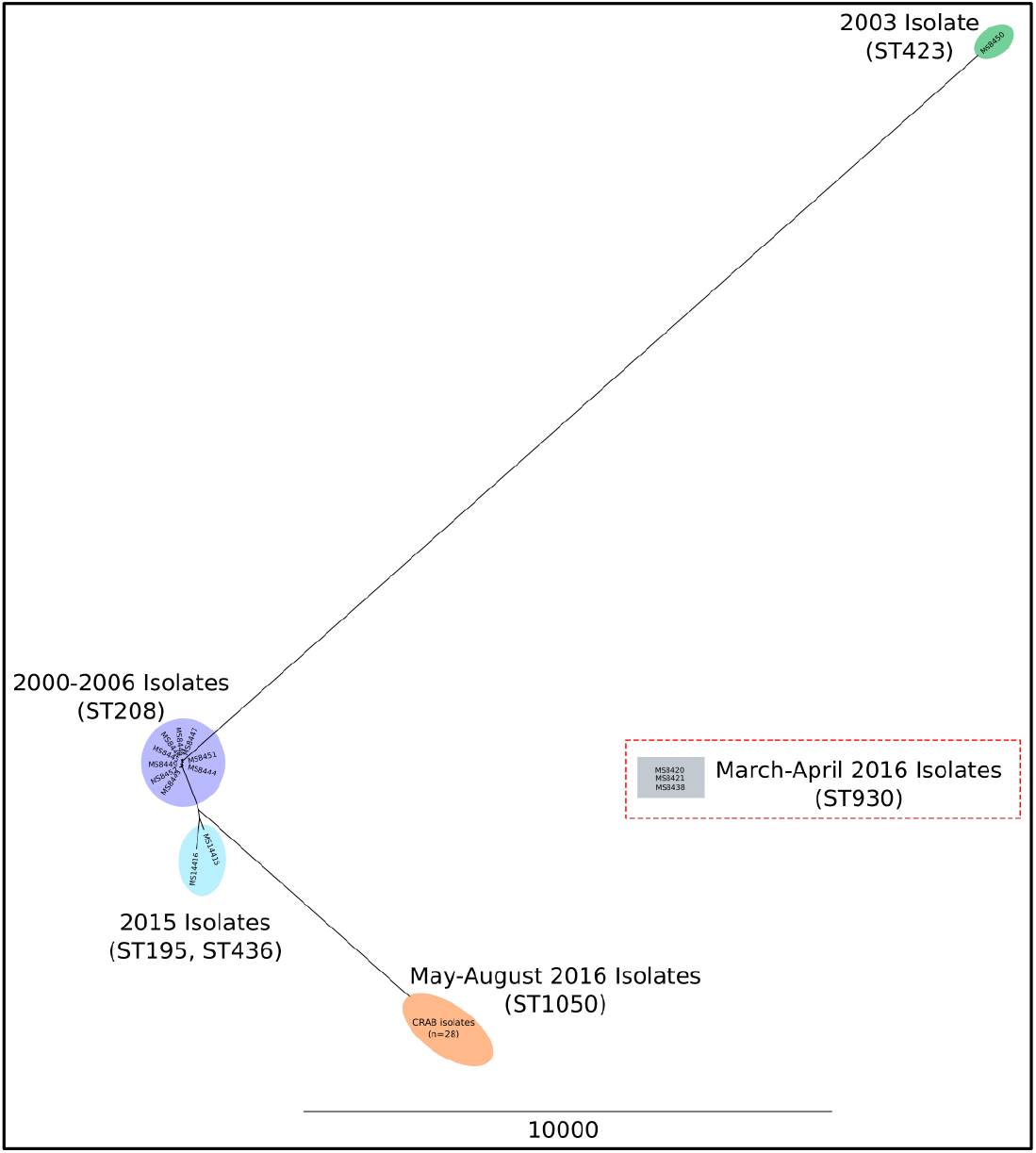
Comparison of outbreak ST1050 CR-Ab isolates to CR-Ab previously isolated from the same hospital between 2000 to early 2016. Tree was created with Parsnp v1.2 (default settings) using the draft *de novo* assemblies for the ST1050 CR-Ab as well as 17 historical CR-Ab from the same hospital. For clarity, the node representing the March-April 2016 isolates (ST930) has been removed from tree as its long branch length obscured the other nodes (see red box inset).

**Supplementary Figure 4:**
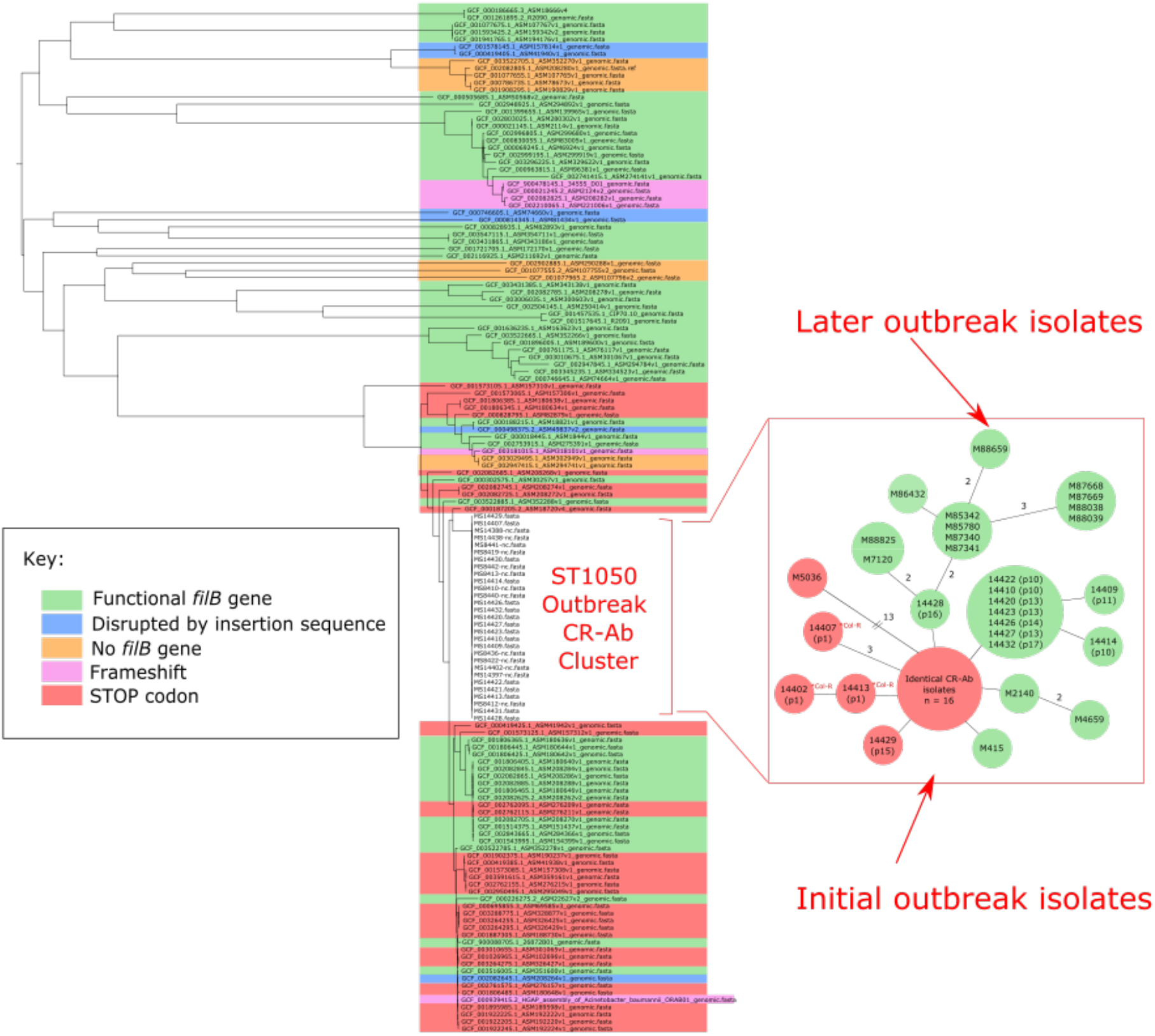
Analysis of nonsense mutation reversion in ST1050 outbreak CR-Ab and complete publicly available. ***A. baumannii* (from NCBI):** Tree built using Parsnp v1.2 (under default settings) with 113 complete *A. baumannii* and the initial outbreak ST1050 CR-Ab genomes (mid-point rooted). Taxa are coloured according to *filB* genotype (refer to key). Inset box shows relationship matrix from Figure 3 (in main text) with nodes coloured according to *filB* genotype. SS17M414 (isolated 3/1/2017) also has a functional *filB* gene, however, is not displayed in the relationship matrix as it clusters in the large group of 16 identical isolates.

**Supplementary Figure 5:**
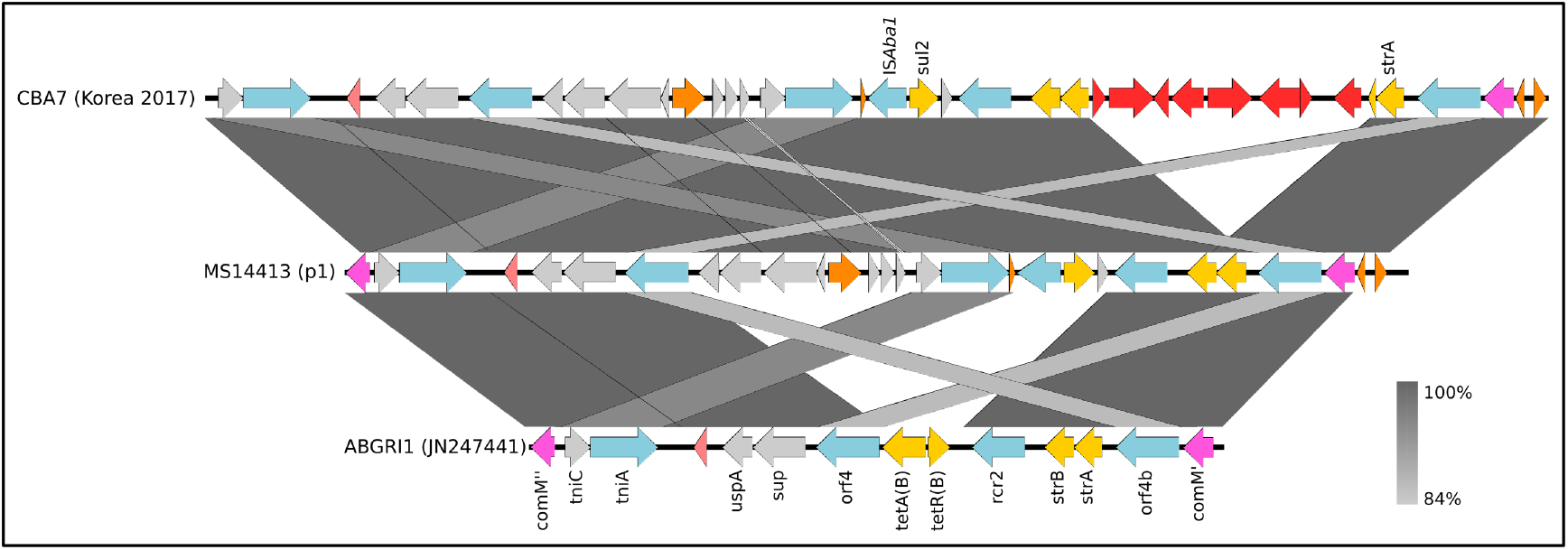
Novel AbGRI1 resistance island in MS14413. BLASTn comparison shown in grey.

**Supplementary Figure 6:**
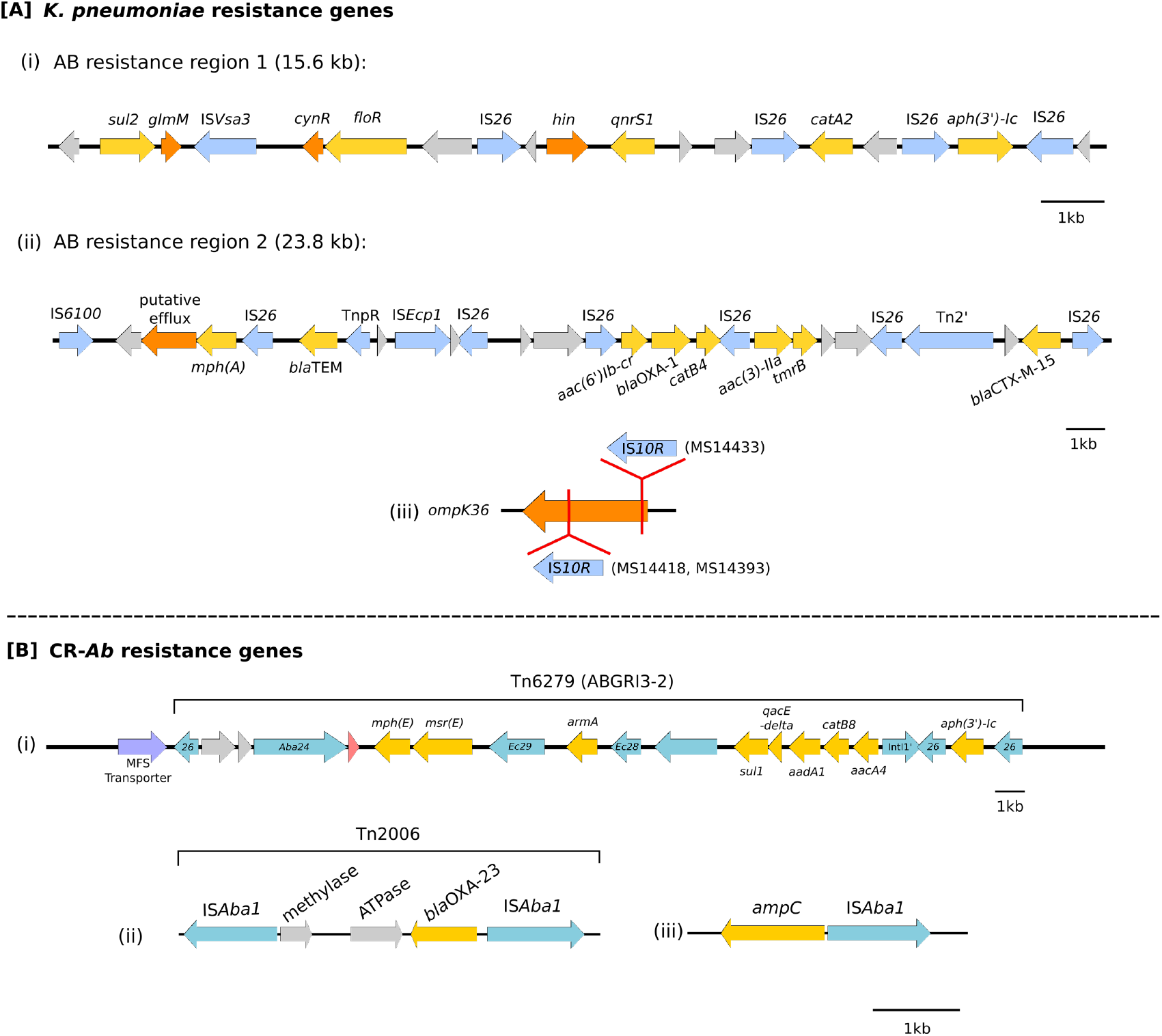
Resistance regions found in K. *pneumoniae* [A] and CR-Ab [B] isolates. arrows represent CDS. Colours represent resistance genes (yellow), regulatory/efflux genes (orange), mobile elements (blue) and hypothetical genes (grey). [A] two resistance regions were found in all *K. pneumoniae* isolates, located on an IncA/C plasmid. [B] CR-Ab isolates were found to have three main mechanisms of resistance: (i) a large transposon Tn6279 (ABGRI3-2), (ii) a smaller transposon (Tn2006) and (iii) an IS*Aba1* element upstream of the intrinsic *ampC* gene.

**Supplementary Figure 7:**
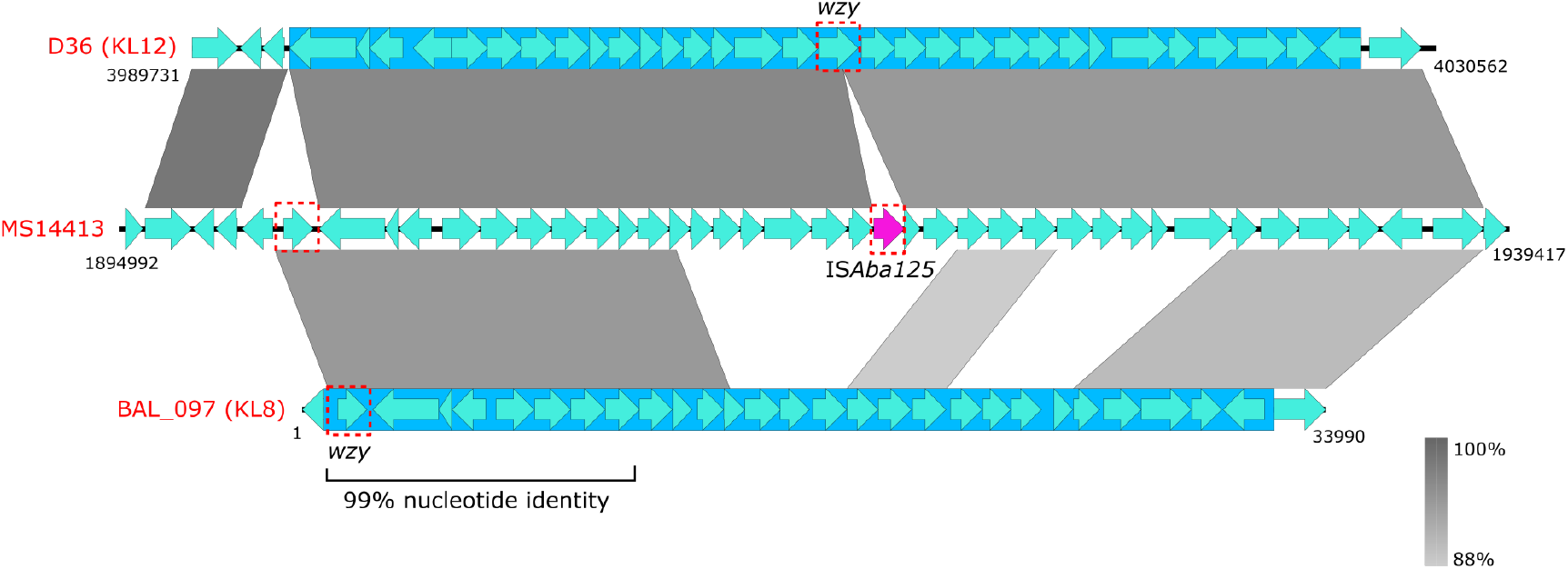
wzy gene positions in capsule (K) region. Light blue arrows represent CDS regions. Dark blue box represents capsule region. Outbreak CR-*Ab* isolate MS14413 was found to have a 97% nucleotide similarity to the KL12 capsule (K) locus found in the GC1 *A. baumannii* strain D36 (GenBank:NZ_CP012952.1) except for an IS*Aba125* insertion sequence in the *wzy* gene. Further comparison found a second *wzy* gene in the same position as in the *A. baumannii* strain BAL_097 (GenBank: KX712116).

**Supplementary Figure 8:**
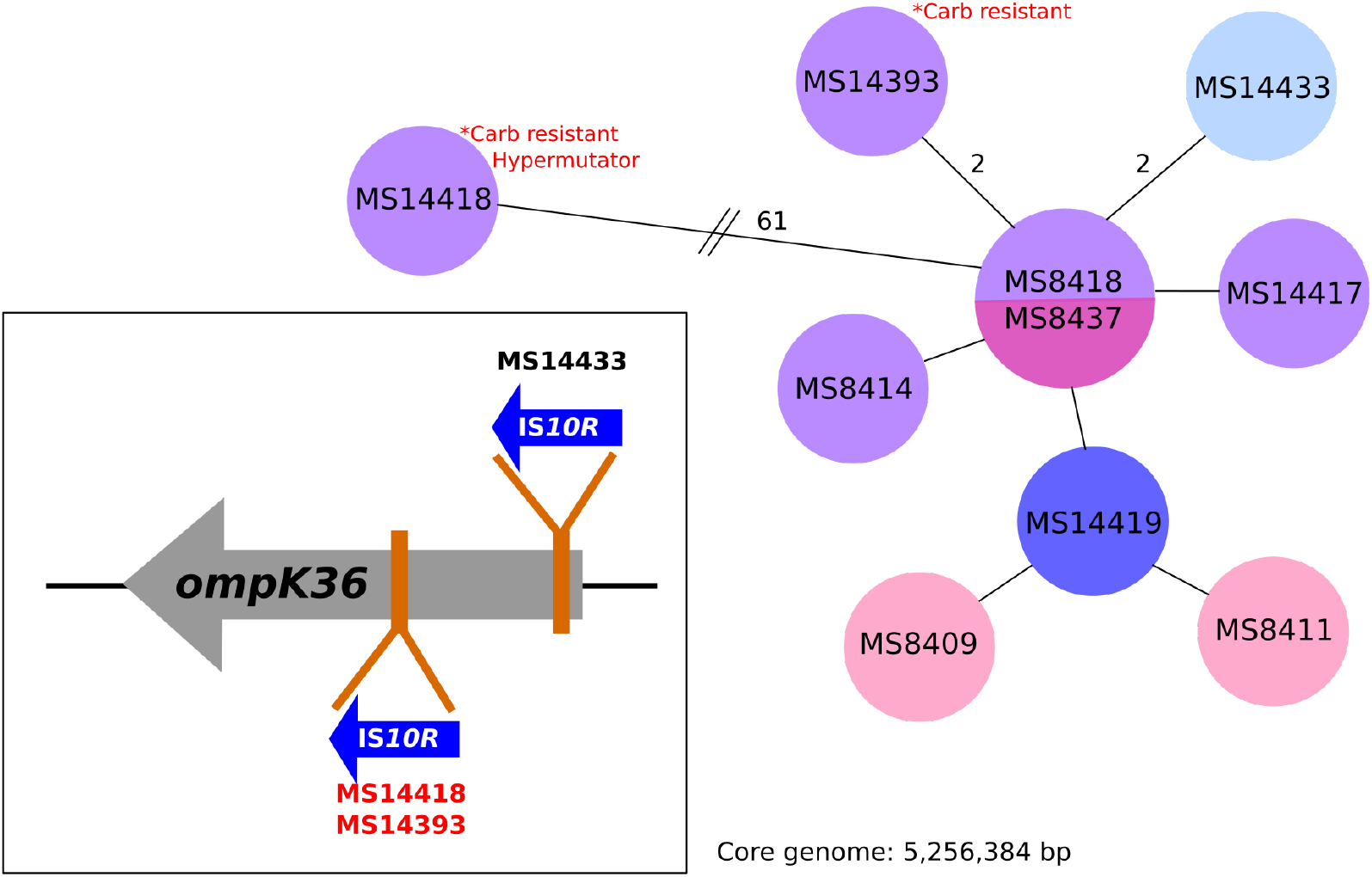
Relationship matrix of ST515. ***Klebsiella pneumoniae* isolates during 2016 outbreak and disruption of *ompK36* outer membrane porin by IS*10R***: coloured circles correspond to patient. Branches represent one SNP difference unless otherwise stated.

**Supplementary Figure 9:**
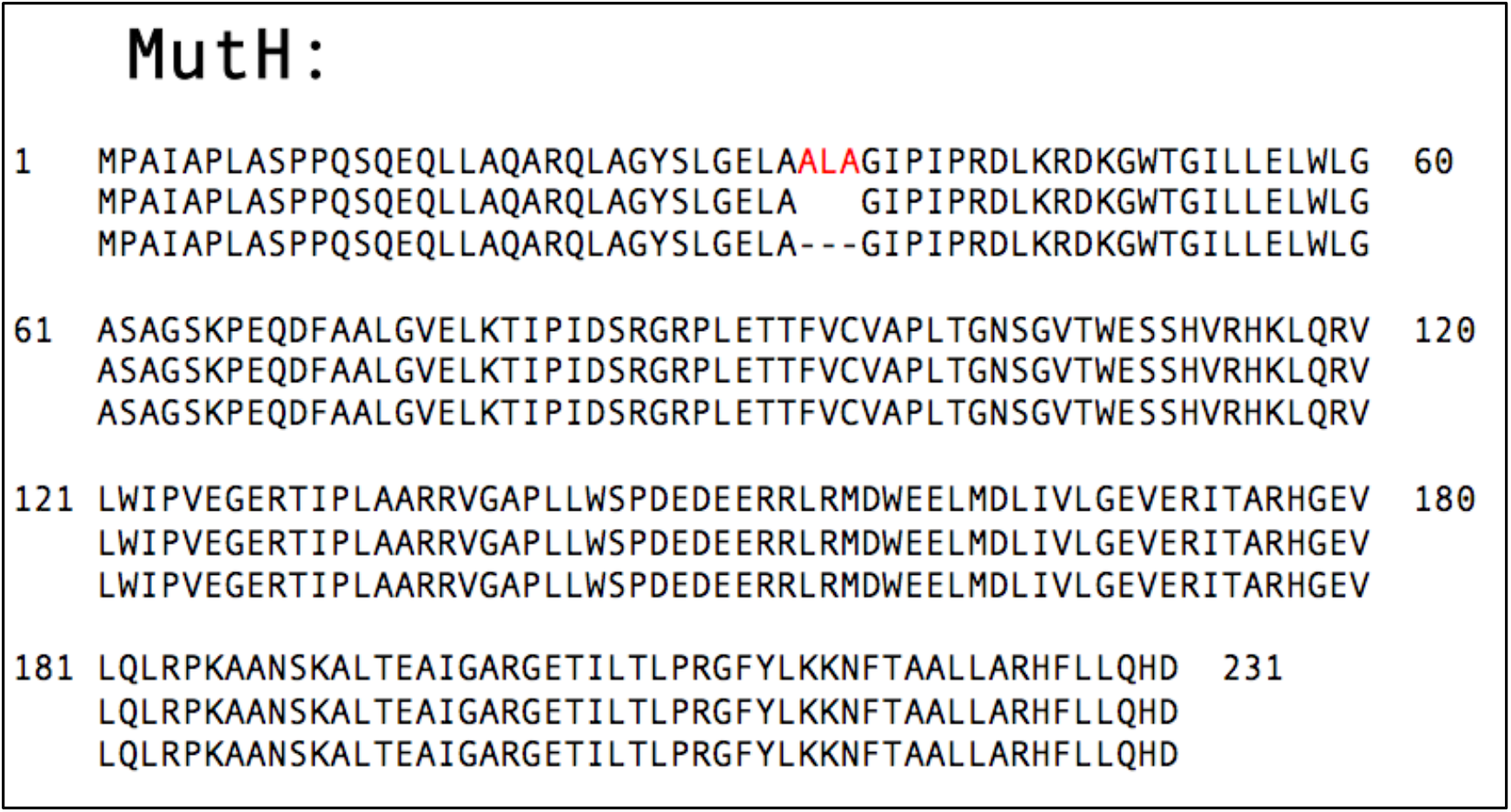
Deletion of three amino acids from MutH protein in MS14418.

**Supplementary Figure 10:**
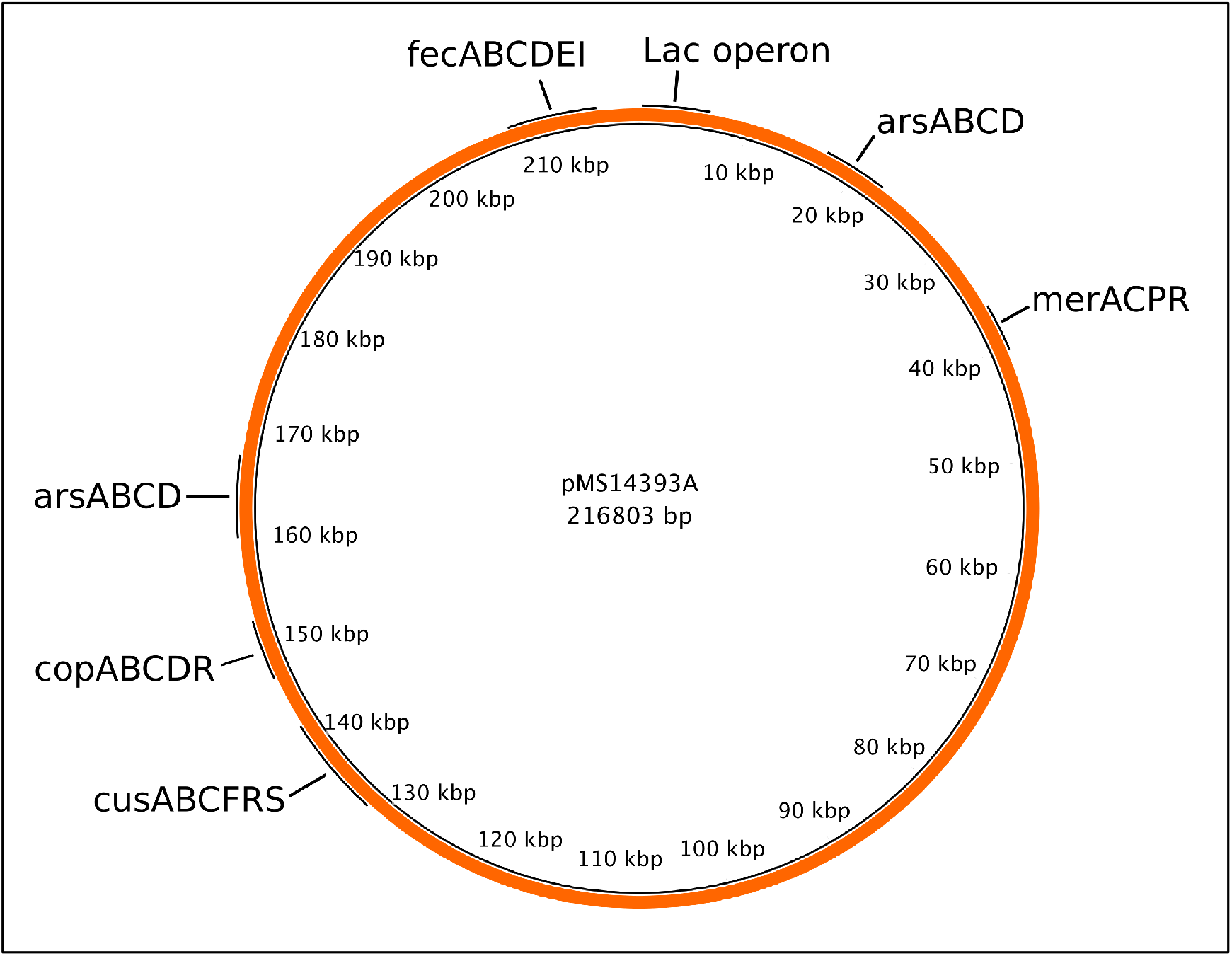
PacBio assembly of IncF plasmid carried by *K. pneumoniae* isolate MS14393.

**Supplementary Figure 11:**
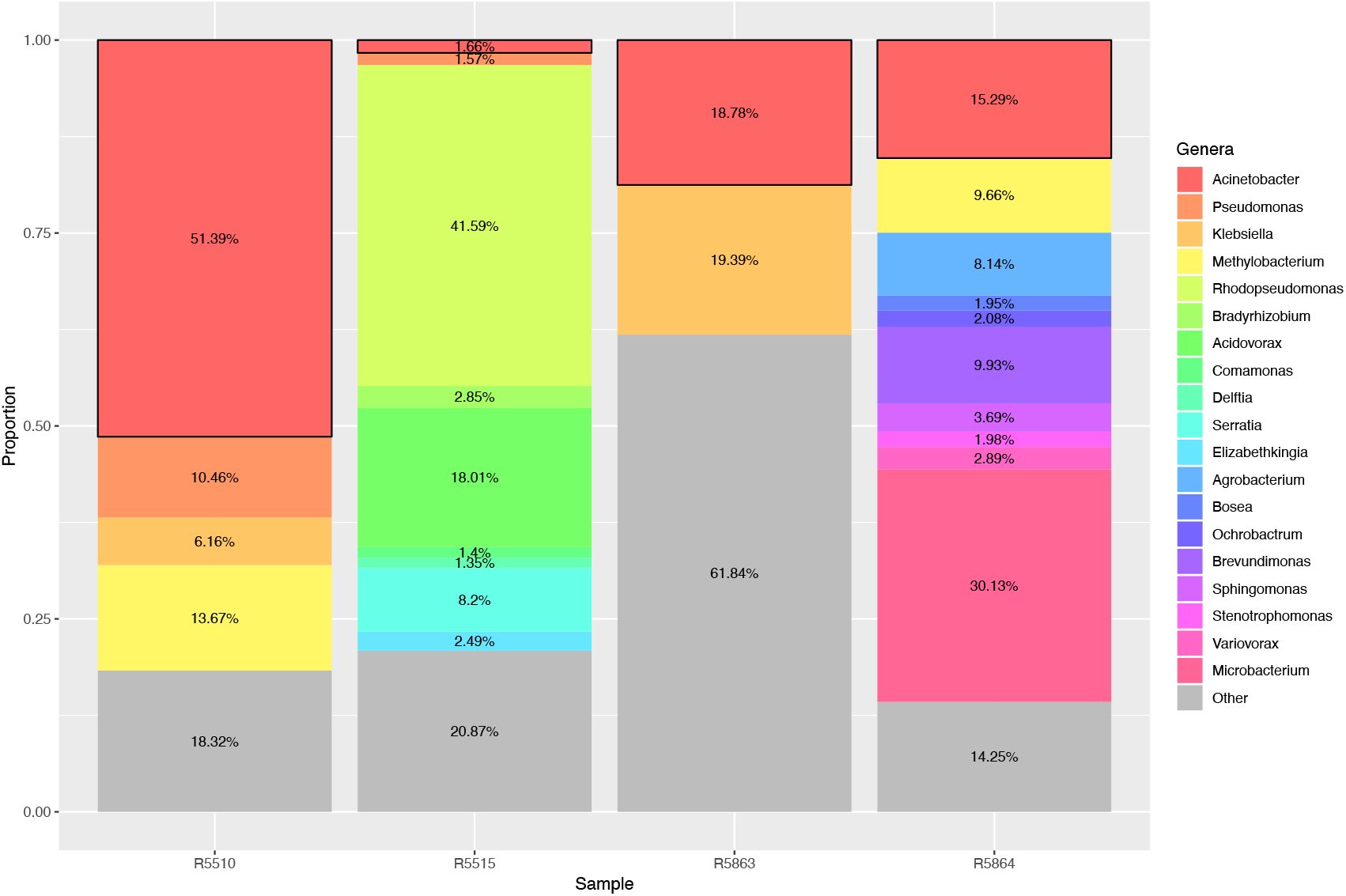
Metagenomic read abundance profiles of environmental surveillance samples. Each column shows the relative abundance of paired-end reads for each environmental sample that were classified at a bacterial genus level by comparing against a database of bacterial genomes from RefSeq. Only bacterial genera with a relative abundance >0.5% are shown as distinct. Genera with an abundance of <0.5% are grouped together as “Other” (grey). Black boxes outlined in black represent abundance of “Acinetobacter”.

**Supplementary Figure 12:**
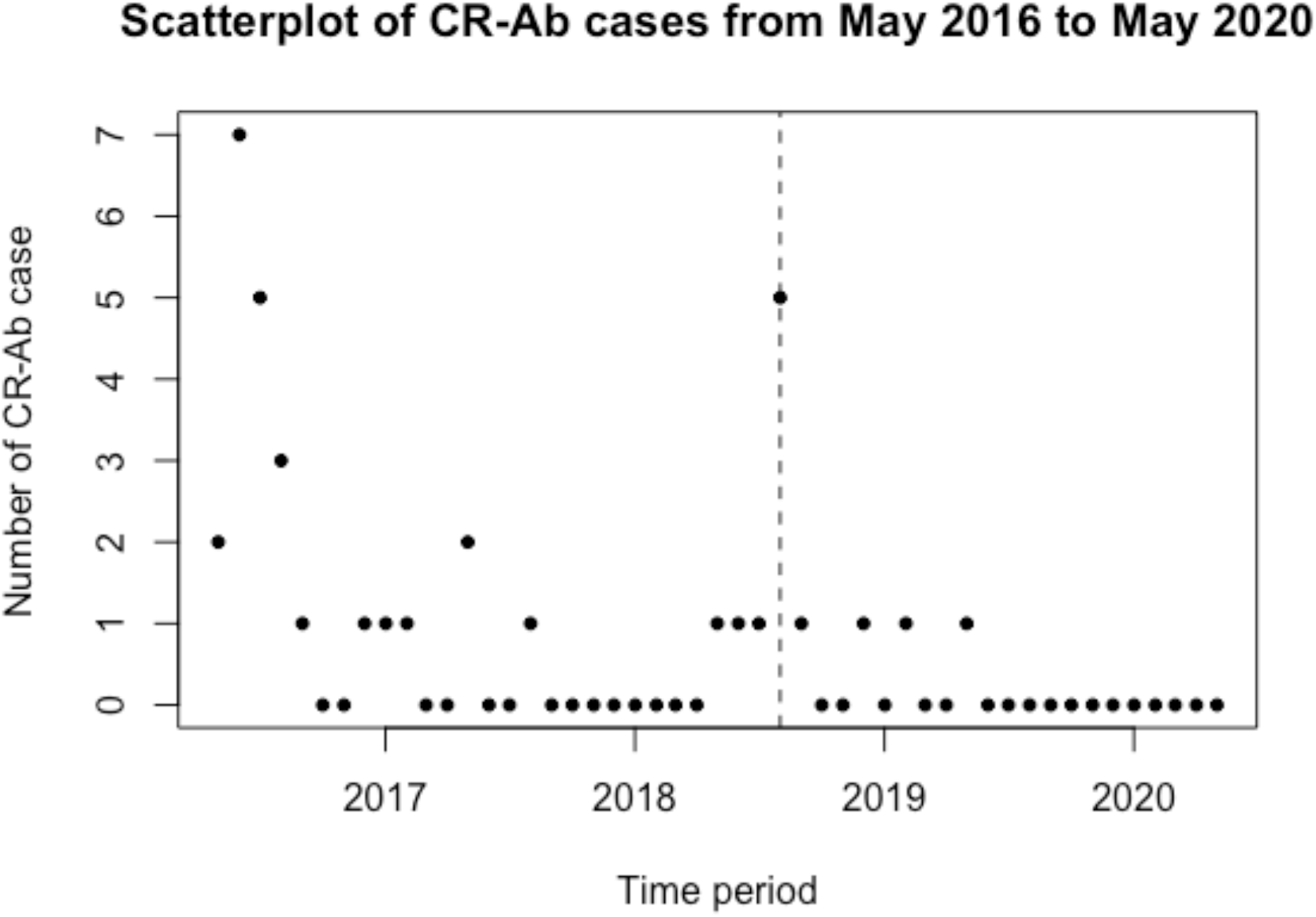
Incidence of ST1050 CR-Ab cases from the time of the initial outbreak until May 2020.

## Notes

### Competing Interest Statement

PNAH has received research grants from MSD, Sandoz and Shionogi Ltd, outside of the submitted work, and speaker fees from Pfizer paid to The University of Queensland. DLP reports receiving grants and personal fees from Shionogi and Merck Sharp and Dohme and personal fees from Pfizer, Achaogen, AstraZeneca, Leo Pharmaceuticals, Bayer, GlaxoSmithKline, Cubist, Venatorx, and Accelerate. JL has received personal fees from Pfizer and MSD and grants from MSD paid to The University of Queensland. The other authors have no conflicts of interest to declare.

## References

1. J. A. Otter et al., Counting the cost of an outbreak of carbapenemase-producing Enterobacteriaceae: an economic evaluation from a hospital perspective. Clin Microbiol Infect 23, 188–196 (2017).

2. M. C. Cruickshank M, Reducing harm to patients from healthcare associated infections: an Australian infection prevention and control model for acute hospitals.. Sydney: Australian Commission on Safety and Quality in Health Care., (2009).

3. C. A. Gilchrist, S. D. Turner, M. F. Riley, W. A. Petri, Jr., E. L. Hewlett, Whole-genome sequencing in outbreak analysis. Clin Microbiol Rev 28, 541–563 (2015).

4. A. Y. Peleg, H. Seifert, D. L. Paterson, Acinetobacter baumannii: emergence of a successful pathogen. Clin Microbiol Rev 21, 538–582 (2008).

5. M. Denton et al., Role of environmental cleaning in controlling an outbreak of Acinetobacter baumannii on a neurosurgical intensive care unit. J Hosp Infect 56, 106–110 (2004).

6. M. Doidge et al., Control of an outbreak of carbapenem-resistant Acinetobacter baumannii in Australia after introduction of environmental cleaning with a commercial oxidizing disinfectant. Infect Control Hosp Epidemiol 31, 418–420 (2010).

7. S. H. Wang et al., Healthcare-associated outbreak due to pan-drug resistant Acinetobacter baumannii in a surgical intensive care unit. J Hosp Infect 53, 97–102 (2003).

8. R. Valencia et al., Nosocomial outbreak of infection with pan-drug-resistant Acinetobacter baumannii in a tertiary care university hospital. Infect Control Hosp Epidemiol 30, 257–263 (2009).

9. M. del Mar Tomas et al., Hospital outbreak caused by a carbapenem-resistant strain of Acinetobacter baumannii: patient prognosis and risk-factors for colonisation and infection. Clin Microbiol Infect 11, 540–546 (2005).

10. J. Nowak et al., High incidence of pandrug-resistant Acinetobacter baumannii isolates collected from patients with ventilator-associated pneumonia in Greece, Italy and Spain as part of the MagicBullet clinical trial. JAntimicrob Chemother 72, 3277–3282 (2017).

11. R. Xie, X. D. Zhang, Q. Zhao, B. Peng, J. Zheng, Analysis of global prevalence of antibiotic resistance in Acinetobacter baumannii infections disclosed a faster increase in OECD countries. Emerg Microbes Infect 7, 31 (2018).

12. C. L. Jones et al., Fatal outbreak of an emerging clone of extensively drug-resistant Acinetobacter baumannii with enhanced virulence. Clin Infect Dis 61, 145–154 (2015).

13. W. Kamolvit, H. E. Sidjabat, D. L. Paterson, Molecular Epidemiology and Mechanisms of Carbapenem Resistance of Acinetobacter spp. in Asia and Oceania. Microb Drug Resist 21, 424–434 (2015).

14. S. Brown, S. Amyes, OXA (beta)-lactamases in Acinetobacter: the story so far. J Antimicrob Chemother 57, 1–3 (2006).

15. L. W. Roberts et al., Integrating multiple genomic technologies to investigate an outbreak of carbapenemase-producing Enterobacter hormaechei. Nat Commun 11, 466 (2020).

16. S. Corvec et al., AmpC cephalosporinase hyperproduction in Acinetobacter baumannii clinical strains. J Antimicrob Chemother 52, 629–635 (2003).

17. S. Quainoo et al., Whole-Genome Sequencing of Bacterial Pathogens: the Future of Nosocomial Outbreak Analysis. Clin Microbiol Rev 30, 1015–1063 (2017).

18. J. C. Kwong et al., Translating genomics into practice for real-time surveillance and response to carbapenemase-producing Enterobacteriaceae: evidence from a complex multi-institutional KPC outbreak. PeerJ 6, e4210 (2018).

19. Y. Jiang et al., The Cost of Responding to an Acinetobacter Outbreak in Critically Ill Surgical Patients. Surg Infect (Larchmt) 17, 58–64 (2016).

20. Z. Sadique, B. Lopman, B. S. Cooper, W. J. Edmunds, Cost-effectiveness of Ward Closure to Control Outbreaks of Norovirus Infection in United Kingdom National Health Service Hospitals. J Infect Dis 213 Suppl 1, S19-26 (2016).

21. D. Marchaim et al., Surveillance cultures and duration of carriage of multidrug-resistant Acinetobacter baumannii. J Clin Microbiol 45, 1551–1555 (2007).

22. A. J. Rodriguez-Acevedo, X. J. Lee, T. M. Elliot, L. G. Gordon, Hospitalization costs for patients colonized with carbapenemase-producing Enterobacterales during an Australian outbreak. J Hosp Infect, (2020).

23. S. R. Harris et al., Evolution of MRSA during hospital transmission and intercontinental spread. Science 327, 469–474 (2010).

24. A. C. Schurch, S. Arredondo-Alonso, R. J. L. Willems, R. V. Goering, Whole genome sequencing options for bacterial strain typing and epidemiologic analysis based on single nucleotide polymorphism versus gene-by-gene-based approaches. Clin Microbiol Infect 24, 350–354 (2018).

25. S. J. Peacock, J. Parkhill, N. M. Brown, Changing the paradigm for hospital outbreak detection by leading with genomic surveillance of nosocomial pathogens. Microbiology 164, 1213–1219 (2018).

26. J. Stimson et al., Beyond the SNP Threshold: Identifying Outbreak Clusters Using Inferred Transmissions. Mol Biol Evol 36, 587–603 (2019).

27. A. Kramer, I. Schwebke, G. Kampf, How long do nosocomial pathogens persist on inanimate surfaces? A systematic review. BMC Infect Dis 6, 130 (2006).

28. A. C. o. S. a. Q. i. H. C. (ACSQHC), “AURA 2019: third Australian report on antimicrobial use and resistance in human health,” (Sydney, 2019).

29. D. E. Wood, S. L. Salzberg, Kraken: ultrafast metagenomic sequence classification using exact alignments. Genome Biol 15, R46 (2014).

30. A. Bankevich et al., SPAdes: a new genome assembly algorithm and its applications to single-cell sequencing. J Comput Biol 19, 455–477 (2012).

31. A. Gurevich, V. Saveliev, N. Vyahhi, G. Tesler, QUAST: quality assessment tool for genome assemblies. Bioinformatics 29, 1072–1075 (2013).

32. T. Seemann, Snippy: fast bacterial variant calling from NGS reads. (2015).

33. T. Seemann, mlst: https://github.com/tseemann/mlst. “This publication made use of the PubMLST website (https://pubmlst.org/) developed by Keith Jolley (Jolley & Maiden 2010, BMC Bioinformatics, 11:595) and sited at the University of Oxford. The development of that website was funded by the Wellcome Trust”.

34. S. G. Bartual et al., Development of a multilocus sequence typing scheme for characterization of clinical isolates of Acinetobacter baumannii. J Clin Microbiol 43, 4382–4390 (2005).

35. T. Seemann, Abricate, Github https://github.com/tseemann/abricate.

36. E. Zankari et al., Identification of acquired antimicrobial resistance genes. J Antimicrob Chemother 67, 2640–2644 (2012).

37. A. Carattoli et al., In silico detection and typing of plasmids using PlasmidFinder and plasmid multilocus sequence typing. Antimicrob Agents Chemother 58, 3895–3903 (2014).

38. M. J. Sullivan, N. K. Petty, S. A. Beatson, Easyfig: a genome comparison visualizer. Bioinformatics 27, 1009–1010 (2011).

39. N. F. Alikhan, N. K. Petty, N. L. Ben Zakour, S. A. Beatson, BLAST Ring Image Generator (BRIG): simple prokaryote genome comparisons. BMC Genomics 12, 402 (2011).

40. A. Rambaut, FigTree. (2009).

41. M. Inouye et al., SRST2: Rapid genomic surveillance for public health and hospital microbiology labs. Genome Med 6, 90 (2014).

42. S. K. Gupta et al., ARG-ANNOT, a new bioinformatic tool to discover antibiotic resistance genes in bacterial genomes. Antimicrob Agents Chemother 58, 212–220 (2014).

43. B. D. Ondov et al., Mash: fast genome and metagenome distance estimation using MinHash. Genome Biol 17, 132 (2016).

44. T. J. Carver et al., ACT: the Artemis Comparison Tool. Bioinformatics 21, 3422–3423 (2005).

45. F. Compain et al., Targeting relaxase genes for classification of the predominant plasmids in Enterobacteriaceae. Int J Med Microbiol 304, 236–242 (2014).

46. S. Koren et al., Canu: scalable and accurate long-read assembly via adaptive k-mer weighting and repeat separation. Genome Res 27, 722–736 (2017).

47. T. Carver, S. R. Harris, M. Berriman, J. Parkhill, J. A. McQuillan, Artemis: an integrated platform for visualization and analysis of high-throughput sequence-based experimental data. Bioinformatics 28, 464–469 (2012).

48. B. J. Walker et al., Pilon: an integrated tool for comprehensive microbial variant detection and genome assembly improvement. PLoS One 9, e112963 (2014).

49. H. Li, R. Durbin, Fast and accurate short read alignment with Burrows-Wheeler transform. Bioinformatics 25, 1754–1760 (2009).

50. T. Seemann, Prokka: rapid prokaryotic genome annotation. Bioinformatics 30, 2068–2069 (2014).

51. A. M. Varani, P. Siguier, E. Gourbeyre, V. Charneau, M. Chandler, ISsaga is an ensemble of web-based methods for high throughput identification and semi-automatic annotation of insertion sequences in prokaryotic genomes. Genome Biol 12, R30 (2011).

52. M. J. Chaisson, G. Tesler, Mapping single molecule sequencing reads using basic local alignment with successive refinement (BLASR): application and theory. BMC Bioinformatics 13, 238 (2012).

53. D. P. Miller, Q. Wang, A. Weinberg, R. J. Lamont, Transcriptome analysis of Porphyromonas gingivalis and Acinetobacter baumannii in polymicrobial communities. Mol Oral Microbiol 33, 364-377 (2018).

54. A. P. Tomaras, C. W. Dorsey, R. E. Edelmann, L. A. Actis, Attachment to and biofilm formation on abiotic surfaces by Acinetobacter baumannii: involvement of a novel chaperone-usher pili assembly system. Microbiology 149, 3473–3484 (2003).

